# Association of [^18^F]Flortaucipir-PET and plasma p-tau217 with tau neuropathology in AD and other neurodegenerative disorders

**DOI:** 10.1101/2025.05.14.25326954

**Authors:** Agathe Vrillon, Salvatore Spina, Jhony Mejia-Perez, Tia Lamore, Claire Yballa, David N. Soleimani-Meigooni, Ganna Blazhenets, Adam L. Boxer, Julio C. Rojas, Argentina L. Lago, William J. Jagust, Bruce L. Miller, Howard J. Rosen, William W. Seeley, Lea T. Grinberg, Gil D. Rabinovici, Lawren Vandevrede, Renaud La Joie

## Abstract

**Background and objectives:** Evaluating tau biomarkers in patients with available autopsy data is critical for validating their sensitivity and specificity to detect Alzheimer’s disease neuropathologic changes (ADNC). We examined the association between tau-PET (using [^18^F]Flortaucipir), plasma ptau-217, and measures of AD neuropathology in a group of clinically-impaired participants from a tertiary dementia center, including patients with AD and FTLD.

**Methods:** We analyzed Flortaucipir-PET (80-100 minutes post-injection acquisition, normalized to inferior cerebellar cortex) from 73 patients with a clinical diagnosis of various neurodegenerative diseases who underwent autopsy at the Neurodegenerative Disease Brain Bank (median [IQR] age: 67 [59, 73] years, 60% male, median [IQR] PET-to-autopsy duration: 3.9 years, [2.1, 5.1]). Standardized uptake value ratios (SUVRs) were extracted from the entorhinal cortex (an early tau region) and the temporal meta-ROI (a widely used AD signature region). A semi-quantitative rating of AD NFT burden in cortical areas was available for 56 participants. Plasma p-tau217 was quantified with Simoa (Janssen®) in 56 participants (median [IQR] PET-to-plasma duration: 1.7 months [0.5, 4.1]).

**Results:** Our cohort included patients with a primary pathological diagnosis of AD (n=39), FTLD (tauopathies n=28, non-tauopathies n=4), and Lewy body dementia (n=2). Flortaucipir SUVRs were elevated in patients with a neuropathological diagnosis of AD compared with non-AD patients. Consistently elevated PET signal was detected in both the entorhinal cortex and temporal meta-ROI of patients with neurofibrillary tangle (NFT) at Braak stage VI and with high ADNC levels. No elevation of Flortaucipir SUVRs was observed at intermediate levels of ADNC. The burden of AD NFTs was correlated with local Flortaucipir SUVRs across cortical brain regions. Plasma p-tau217 concentrations were increased in patients at Braak stage V and VI and strongly correlated with Flortaucipir SUVRs across ROIs (r’s≥0.75), driven by Braak V-VI cases. Flortaucipir SUVRs and plasma p-tau217 concentrations identified high Braak stages (V-VI) and high/intermediate ADNC levels with similarly high performance (area under the curve≥ 0.90).

**Discussion:** Flortaucipir-PET and plasma p-tau217 both displayed strong specificity for primary AD neuropathological diagnosis but lacked sensitivity to detect early AD-related tau co-pathology in patients with a non-AD diagnosis.

## INTRODUCTION

The detection of Alzheimer’s disease (AD) related tau pathology in the brain is now possible during life with tau-specific biomarkers. Positron emission tomography (PET) ligands binding to aggregated tau protein, including [^18^F]Flortaucipir, have demonstrated the ability to identify tau pathology associated with AD in vivo^1,2^. PET-to-autopsy studies show Flortaucipir’s high performance in detecting advanced tau neuropathology, i.e., neurofibrillary tangles (NFT) Braak stages V and VI. In that context, Flortaucipir-PET has been approved by the FDA for the detection of NFT pathology in patients being evaluated for AD^3^. However, Flortaucipir-PET’s capacity to detect more moderate AD neuropathological changes (ADNC), corresponding to intermediate NFT Braak stages, remains uncertain. Autopsy studies have shown inconsistent tracer uptake at NFT Braak stages III and IV^2,4^. Importantly, Flortaucipir-PET has also been used in patients with non-AD dementia, including primary tauopathies and non-tauopathy diseases^5–7^. Studies report mixed results with none-to-mild tracer uptake, below the signal intensity observed in AD and in non-AD tauopathies, with tracer retention overlapping with off-target binding regions.

In addition to imaging, plasma phosphorylated-tau (p-tau) isoforms have emerged as promising non-invasive biomarkers for AD diagnosis^8^. Among those, p-tau217 has outperformed all other plasma biomarkers to detect AD neuropathology and to discriminate AD from non-AD causes of cognitive impairment^9,10^. In autopsy-confirmed cohorts, plasma p-tau217 concentrations have been associated with both amyloid-β (Aβ) and tau pathologies^11^. Plasma p-tau217 has also demonstrated a high agreement with tau PET^12–14^.

While Flortaucipir-PET and plasma p-tau217 have demonstrated strong performance in detecting AD neuropathology, they have mostly been evaluated in separate studies, and head-to-head comparisons are lacking. Thus, our aims were to study i/ the concordance of tau pathology measured through the NFT Braak stage with Flortaucipir-PET findings across a wide range of neurodegenerative disorders, ii/ the association with semi-quantitative measures of AD NFT burden, iii/ the association of PET and autopsy findings with the newly available plasma p-tau217 marker.

## METHODS

### Cohort

We systematically included all patients from the UCSF Neurodegenerative Diseases Brain Bank (NDBB) of the Memory and Aging Center/Alzheimer’s Disease Research Center who underwent *antemortem* Flortaucipir-PET and *postmortem* examination between 2013 and 2024. Twenty of those cases were described in Soleimani-Meigooni *et al*^5^. All patients underwent comprehensive history gathering, neurological evaluation, neuropsychological assessment, structural neuroimaging, and genetic testing for diagnosis (detailed in **e-Methods)**.

An independent, non-autopsy control group was included to compute w-maps (age and sex-adjusted comparisons of individual images to controls) and to define regional Flortaucipir-PET thresholds for tau positivity for our regions of interest. The group consisted of 103 cognitively unimpaired amyloid-PET-negative individuals who had undergone Flortaucipir-PET (male: 47%, mean age 67.2±16.4) from the Berkeley Aging Cohort Study. A stringent global PiB DVR threshold of 1.065 (corresponding to 10 Centiloids^15^) was used to determine Aβ positivity to rule out the presence of detectable amyloidosis, consistent with recent guidelines^16^.

### Neuropathology

The neuropathological analysis was performed according to the NDBB standardized sampling and staining protocols^17^. All cases were reviewed by a board of neuropathologists (L.T.G., S.S., and W.W.S.), and a consensus diagnosis was achieved.

Neuropathological diagnoses were based on consensus criteria for AD, progressive supranuclear palsy (PSP), corticobasal degeneration (CBD), frontotemporal lobar degeneration (FTLD) associated with *MAPT* mutation (FTLD-MAPT), FTLD with motor neuron disease associated with *FUS* mutation (FTLD-FUS), Pick’s disease, FTLD with TDP-43 inclusions (FTLD-TDP), argyrophilic grain disease (AGD), Lewy body disease (LBD), and chronic traumatic encephalopathy (CTE)^18–25^. The “primary” neuropathological diagnosis was the pathological entity whose severity and topographical distribution were considered to drive the clinical presentation of the disease.

AD neuropathological changes (ADNC) were assessed using the NIA-AA guidelines as “None,” “Low,” “Intermediate,” or “High” ADNC. This included scoring of Thal phase, Braak and Braak staging system for neurofibrillary tangles (NFT Braak), and Consortium to Establish a Registry for AD (CERAD) scale for neuritic plaques^25–27^. Primary AD neuropathological diagnosis was defined as intermediate or high ADNC, considered as the pathology that was most likely to explain the clinical syndrome^18^.

In addition to NFT Braak staging, a semi-quantitative measurement of AD NFT pathology was performed across cortical regions^28^. The burden of AD NFT was assessed by a visual rating using a four-point scale (absent: 0, mild:1, moderate: 2, or severe: 3) on slides immunostained with CP13 antibody. Assessed brain regions included the entorhinal cortex, CA1/subiculum, CA2, CA3/CA4, dentate gyrus, amygdala, inferior temporal, anterior middle cingulate, middle frontal, angular, precentral, postcentral, and calcarine cortices. The average AD NFT burden in each sampled region was matched to the average of the PET-SUVR of the corresponding bilateral ROI. We averaged the scores of all regions into an average burden score that was correlated to the global tau SUVR and plasma p-tau217 concentrations.

### Imaging

#### Image acquisition

Patients and controls underwent structural MRI on a 3T scanner to acquire T1-weighted magnetization-prepared rapid gradient-echo (MPRAGE) sequences for PET processing (**eMethods**). PET scanning was performed at the Lawrence Berkeley National Laboratory on a Siemens Biograph PET/CT scanner (n=69 patients and all controls) and at UCSF China Basin on a GE Discovery PET/CT scanner (n=4 patients). Flortaucipir PET images were acquired 80 to 100 minutes (four, 5-minute frames) after injection of 10 mCi of radiotracer. Head CT scans were used for attenuation correction. All images were smoothed to a final resolution of 6.5 x 6.5 x 7.25 mm full width at half maximum.

#### Image processing

Processing of Flortaucipir images was performed using Statistical Parametric Mapping 12 (SPM, Wellcome Department of Imaging Neuroscience, Institute of Neurology, London, UK) and Freesurfer version 7 (surfer.nmr.mgh.harvard.edu). Flortaucipir-PET frames were realigned, averaged, and coregistered to the corresponding T1 MRI images. Standardized uptake value ratio (SUVR) images were created using inferior cerebellar gray matter as the reference region. Individual SUVR images are presented in **Supp. Figure 1**. FreeSurfer was used to define cortical regions of interest (ROI) in native subject space, according to the Desikan_Killiany atlas^29^. The mean SUVR value was extracted from each ROI. The Pick Atlas toolbox was used to define basal ganglia ROIs (**eMethods**). For each case, w-maps were created to display voxel-wise z-scores in reference to the control groups, adjusting for age and sex (**Supp. Figure 1, eMethods**).

We reproduced our main analyses after applying partial volume correction (PVC), with a Geometric Transfer Matrix approach using the PETPVC toolbox^30^.

#### Regions of interest

Key ROIs included the entorhinal cortex as an early tau region and the temporal meta-ROI (which includes the entorhinal cortex, amygdala, inferior/middle temporal gyri, fusiform gyrus, and parahippocampal gyrus) that is a validated AD summary region^31^. We derived independent SUVR thresholds for positivity using the non-autopsy control group to contrast clinically derived cut points to the pathological gold standard. We defined the threshold for positivity as the mean + 2 standard deviations above our control cohort, yielding cut points of 1.39 for the entorhinal ROI and 1.35 for the temporal meta-ROI. Using the 95th percentile value from the control group yielded similar thresholds (entorhinal ROI: 1.37; temporal meta-ROI: 1.28) that did not impact any of the conclusions (**Supp. Figure 2**). A global tau ROI combining all Braak regions (corresponding to the full cortex and the amygdala) was used to explore the correlation with semi-quantitative pathological measurements.

#### Tau-PET visual read

Flortaucipir scans were visually read according to FDA-approved guidelines^3^. Elevated tracer uptake in posterolateral temporal, occipital, or parietal/precuneus regions in either hemisphere, with or without frontal involvement, defined a positive visual read. The absence of tracer uptake, or tracer uptake limited to medial temporal, anterolateral temporal, and/or frontal regions, resulted in a negative visual read. Isolated small foci of signal, white matter uptake, and binding in areas of unspecific off-target binding (basal ganglia, brainstem, superior cerebellum) were not defined as tau positive.

#### Amyloid PET

Amyloid PET was available for most participants (70/73) from the autopsy sample using [^11^C]Pittsburgh compound B (PIB; n=66), [^18^F]Florbetapir (n=3), or [^18^F]Florbetaben (n=1) at a median of 0 (0.0, 1.3) months from tau PET. Aβ-PET positivity was based on visual interpretation (**eMethods**).

### Plasma p-tau217

Plasma p-tau217 measurement was available for a subset of 56 patients, which was obtained near the time of PET acquisition (median [IQR] plasma-to-PET duration: 1.6 [0.24, 3.7] months). Plasma p-tau217 measurements were not available for control participants. These measurements were performed in the context of a larger plasma biomarker study reported by Vandevrede *et al*^10^. Blood samples were obtained by venipuncture in EDTA tubes and stored at −80 °C until analysis (on the 1^st^ thawing). Plasma p-tau217 concentrations were measured in duplicate using a single-molecule array (Simoa) assay^32,33^. The coefficients of variation were below 5% overall.

### Statistical analysis

Analyses were performed using R version 4.3.2 and Graphpad Prism. We reported demographics and pathological scores as median (interquartile interval) and frequency (percent) by diagnosis when the sample size was ≥3. Otherwise, we reported individual values. Plasma p-tau217 concentrations were log-transformed prior to analysis. SUVR and semi-quantitative AD NFT burden association were explored using Spearman’s rank correlations. The association between plasma p-tau217 concentrations and Flortaucipir-PET SUVRs was investigated using Spearman’s rank correlations in the whole cohort and in the AD group. The area under the curve (AUC) derived from receiver operating characteristic (ROC) curves was used to explore the performance of the Flortaucipir-PET ROIs and plasma p-tau217 concentrations to identify ADNC levels and NFT Braak stages. The analysis was performed in the 56 participants with both Flortaucipir-PET and p-tau217 available. Age, sex, and plasma/PET-to-autopsy delay were included as covariates. AUCs were compared using DeLong tests^34^. Overall, p-values ≤ 0.05 were considered significant.

### Standard Protocol Approvals, Registrations, and Patient Consents

Written informed consent was obtained from all participants at the time of recruitment. The study was approved by institutional review boards at UCSF, University of California Berkeley, and Lawrence Berkeley National Laboratory (LBNL).

### Data availability

Qualified researchers from academic, not-for-profit institutions can request de-identified data associated with this study through the UCSF Memory and Aging Center (https://memory.ucsf.edu/research-trials/professional/open-science) after obtaining IRB approval from the UCSF Human Research Protection Program.

## RESULTS

Participants’ characteristics are summarized in **Table 1**. The cohort consisted of 73 patients (median age, 67 [59, 73] years; sex, 60% male; median PET-to-autopsy delay, 3.9 [2.1, 5.1] years), including 39 cases with a pathological diagnosis of AD and 34 cases with a pathological diagnosis of non-AD dementia.

**Table 1.** Demographics and AD neuropathologic scores. Categorical variables are presented as median [Q1, Q3] for groups of n≥3. Individual values are presented for groups of n=1-2. APOE4 carriership is presented as the number of carriers/number of cases tested (% of positivity). *Plasma p-tau217 measurements were available for a subsample of n=56 participants, which is described in Supp.Table 3. To limit identifying information, ages are presented as age range for diagnosis group of n=1 to 2. Abbreviations: ADNC, Alzheimer’s disease neuropathologic change; AGD, argyrophilic grain disease; CBD, corticobasal degeneration; CERAD, Consortium to Establish a Registry for Alzheimer’s Disease; CTE, chronic traumatic encephalopathy; FTLD, frontotemporal lobar degeneration; Fus, RNA-binding protein fused in sarcoma; LBD, Lewy body dementia; MMSE, mini-mental state examination; NFT, neurofibrillary tangles; PSP, progressive supranuclear palsy.

|  | Overall | AD | PSP | CBD | CTE | FTLD-<br>MAPT | Pick's | AGD | FTLD<br>TDP-43 | FTLD-FUS | LBD |
| --- | --- | --- | --- | --- | --- | --- | --- | --- | --- | --- | --- |
|  | N=73 | N=39 | N=11 | N=9 | N=2 | N=3 | N=2 | N=1 | N=3 | N=1 | N=2 |
| Age at PET | 67 [59, 73] | 66 [59, 73] | 72 [67, 75] | 63 [57, 71] | 66-70, 76-80 | 45 [41, 56] | 61-65, 71-75 | 65-70 | 54 [51, 61] | 31-40 | 71-75, 81-85 |
| Sex, male | 44 (60%) | 18 (46%) | 9 (82%) | 6 (67%) | 2 (100%) | 2 (67%) | 2 (100%) | 1 (100%) | 1 (33%) | 1 (100%) | 2 (100%) |
| ApoE4<br>carriership | 31/67 (45%) | 22/37 (59%) | 4/9 (44%) | 2/9 (22%) | 0/2 (0%) | 1/1(50%) | 1/2 (50%) | 0/1 (0%) | 1/3 (33%) | 0 /1 (0%) | 0/2 (0%) |
| MMSE | 22 [16, 26] | 19 [15, 24] | 26 [24, 27] | 27 [17, 29] | 27, 22 | 5 [4, 17] | 25, 28 | 29 | 24 [23, 25] | 22 | 0, 28 |
| Plasma<br>p-tau217<br>pg/mL* | 0.155 [0.089,<br>0.309] | 0.312 [0.211,<br>0.437] | 0.067 [0.046,<br>0.091] | 0.130 [0.093,<br>0.145] | 0.103 / na | 0.112 [0.103,<br>0.121] | 0.0517 / na | 0.0993 | 0.042 [0.034,<br>0.057] | 0.0358 | 0.242, 0.238 |
| Missing | 17 (23%) | 12 (31%) | 1 (9%) | 1 (11%) | 1 (50%) | 1 (33%) | 1 (50%) | - | - | - | - |
| Age at autopsy | 70 [64, 77] | 70 [64, 78] | 75 [70, 77] | 65 [64, 76] | 71-75, 81-85 | 53 [46, 62] | 66-70, 76-80 | 66-70 | 58 [54, 64] | 31-40 | 71-75, 86-90 |
| Pet-to-autopsy<br>delay, years | 3.9 [2.1, 5.1] | 4.4 [3.3, 5.5] | 2.0 [1.0, 3.8] | 2.6 [2.1, 4.5] | 4.3, 6.0 | 3.0 [2.6, 5.6] | 3.0, 4.7 | 0.32 | 1.7 [1.6, 3.0] | 1.0 | 1.9, 2.9 |
| Thal phase |  |  |  |  |  |  |  |  |  |  |  |
| 0 | 10 (14%) | - | 2 (18%) | 3 (33%) | 1 (50%) | 1 (33%) | - | 1 (100%) | 1 (33%) | 1 (100%) | - |
| 1 | 5 (7%) | - | 2 (18%) | - | - | - | - | - | 2 (67%) | - | 1 (50%) |
| 2 | 9 (12%) | - | 2 (18%) | 4 (44%) | - | 1 (33%) | 2 (100%) | - | - | - | - |
| 3 | 3 (4%) | 1 (3%) | 2 (18%) | - | - | - | - | - | - | - | - |
| 4 | 5 (7%) | - | 2 (18%) | 1 (11%) | 1 (50%) | - | - | - | - | - | 1 (50%) |
| 5 | 40 (55%) | 38 (97%) | 1 (9%) | - | - | 1 (33%) | - | - | - | - | - |
| Unstaggable | 1 (1%) | - | - | 1 (11%) | - | - | - | - | - | - | - |
| NFT Braak stage |  |  |  |  |  |  |  |  |  |  |  |
| 0 | 4 (5%) | - | - | - | - | 2 (67%) | 1 (50.0%) | - | - | 1 (100%) | - |
| 1 | 10 (14%) | - | 3 (27%) | 2 (22%) | - | - | 1 (50.0%) | - | 3 (100%) | - | 1 (50.0%) |
| 2 | 7 (10%) | - | 4 (36%) | 3 (33%) | - | - | - | - | - | - | - |
| 3 | 7 (10%) | - | 2 (18%) | 4 (44%) | - | - | - | 1 (100%) | - | - | - |
| 4 | 3 (4%) | - | 2 (18%) | - | - | 1 (33%) | - | - | - | - | - |
| 5 | 3 (4%) | 2 (5%) | - | - | - | - | - | - | - | - | 1 (50.0%) |
| 6 | 37 (51%) | 37 (95%) | - | - | - | - | - | - | - | - | - |
| Non conformant | 2 (3%) | - | - | - | 2 (100%) | - | - | - | - | - | - |
| **AD CERAD** |  |  |  |  |  |  |  |  |  |  |  |
| None | 19 (26%) | - | 5 (45%) | 4 (44%) | 1 (50%) | 2 (67%) | 1 (50%) | 1 (100%) | 3 (100%) | 1 (100%) | 1 (50%) |
| Sparse | 3 (4%) | - | 1 (9%) | 1 (11%) | - | - | 1 (50%) | - | - | - | - |
| Moderate | 8 (11%) | - | 4 (36%) | 3 (33%) | - | 1 (33%) | - | - | - | - | - |
| Frequent | 43 (59%) | 39 (100%) | 1 (9%) | 1 (11%) | 1 (50%) | - | - | - | - | - | 1 (50%) |
| **ADNC** |  |  |  |  |  |  |  |  |  |  |  |
| Not | 10 (14%) | - | 2 (18%) | 3 (33%) | 1 (50%) | 1 (33%) | - | 1 (100%) | 1 (33%) | 1 (100%) | - |
| Low | 15 (20%) | - | 6 (55%) | 3 (33%) | - | 1 (33%) | 2 (100%) | - | 2 (67%) | - | 1 (50%) |
| Intermediate | 7 (10%) | 1 (3%) | 3 (27%) | 2 (22%) | - | 1 (33%) | - | - | - | - | - |
| High | 39 (53%) | 38 (97%) | - | - | - | - | - | - | - | - | 1 (50%) |
| Unstagnable | 2 (3%) | - | - | 1 (11%) | 1 (50%) | - | - | - | - | - | - |

All participants with AD as a primary pathology had dementia at the time of death, except one case who had mild cognitive impairment. A positive amyloid PET scan was observed for all 39 AD cases (100%). AD cases had a Braak stage of VI for 37 cases (95%) and V for 2 cases (5%) and high levels of ADNC, except for one case with intermediate levels.

Non-AD cases had diagnoses of 4-repeat (4R) tauopathies such as PSP (n=11), CBD (n=9), FTLD-*MAPT* (n=3), and AGD (n=1), or 3-repeat (3R) tauopathy Pick’s disease (n=2), or CTE, a mixed 3R/4R tauopathy (n=2), while non-tauopathy cases included FTLD-TDP cases (n=3), FTLD-*FUS* (n=1), and LBD (n=2). Genetic statuses are detailed in **eMethods**. In 32 non-AD cases with available amyloid PET, 6 (22%) were positive (PSP n=2, CBD n=2, FTLD-MAPT n=1, CTE n=1, and LBD n=1). CTE cases were not given an NFT Braak stage because it is a mixed 3R/4R tauopathy, similar to AD, which limits the attribution of NFT pathology to AD or CTE. Of the 34 cases with a non-AD primary neuropathological diagnosis, 21 (62%) were Braak stage 0-II, 10 (29%) III-IV and 1 (3%) Braak V. One case (3%) had high levels of comorbid ADNC (primary neuropathology: LBD), 6 (18%) had intermediate levels of ADNC (primary neuropathologies: PSP n=3, CBD n=2 and FTLD-MAPT n=1), and 15 (44%) had low ADNC levels.

### Flortaucipir-PET across neuropathological diagnoses

Patients with primary AD neuropathology displayed elevated entorhinal Flortaucipir SUVRs compared with non-autopsy controls, with 37/39 (95%) cases above the positivity threshold defined using the non-autopsy controls (**Figure 1A**). Using the temporal meta-ROI, all AD cases but one case (Braak V) displayed increased SUVRs above the positivity cutoff (**Figure 1B**). Non-AD cases had SUVR values in range with controls, below the positivity threshold for both ROIs. Similar findings were found across all six Braak stage regions (**Supp. Figure 3**) or when using partial volume-corrected PET data (**Supp. Figure 4**).

**Figure 1.**
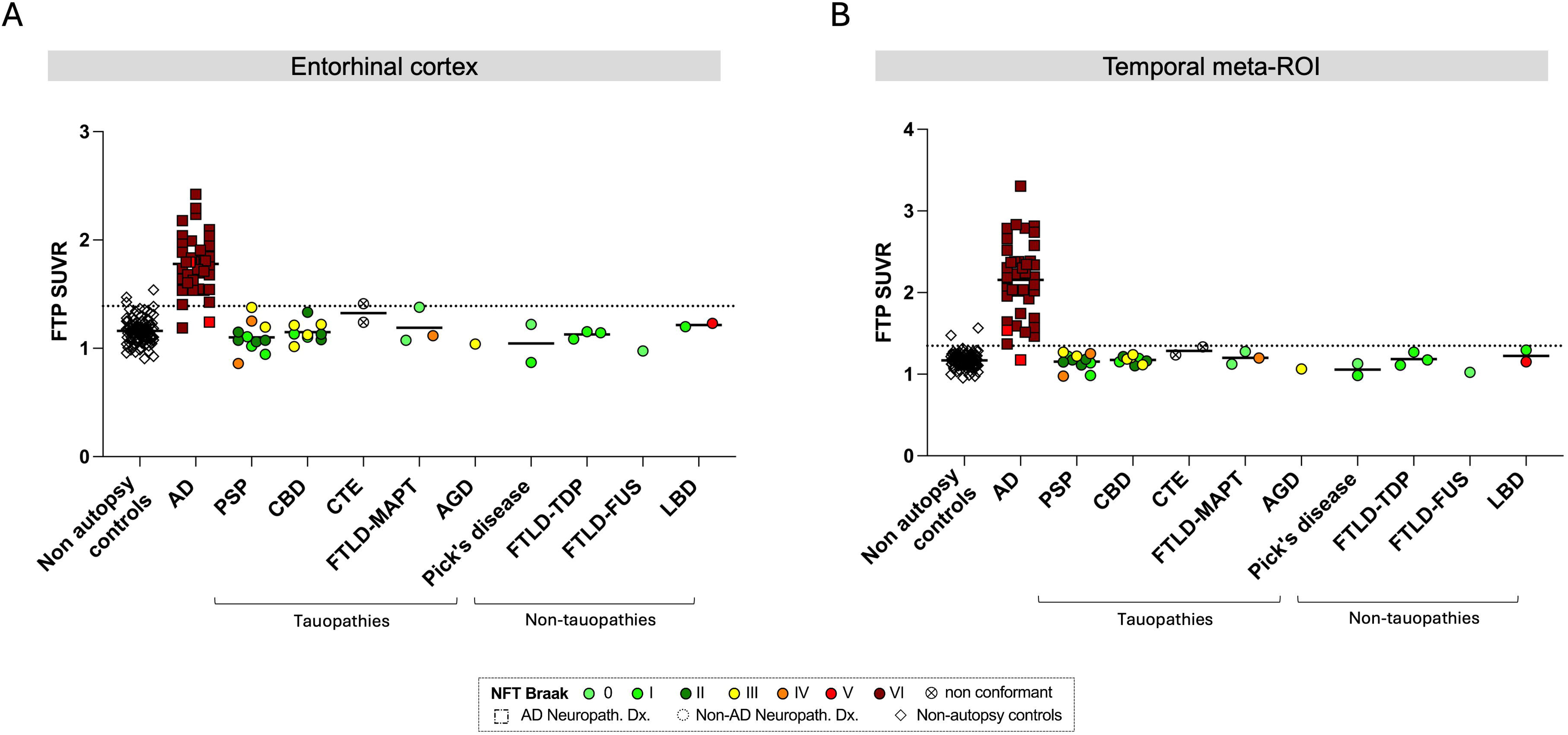
Association of Flortaucipir SUVRs with neuropathological diagnoses. **A,** Entorhinal FTP SUVR by neuropathologic diagnosis. **B,** Temporal meta-ROI SUVR by neuropathologic diagnosis. The horizontal line represents the cut-off for SUVR positivity derived from the non-autopsy control group (mean + 2SD). Individual points are color-coded by their NFT Braak stage. Abbreviations: AGD, argyrophilic grain disease; CBD, corticobasal degeneration; CTE, chronic traumatic encephalopathy; FTLD-*FUS*, frontotemporal lobar degeneration due to a *FUS* mutation; FTLD-*MAPT*, frontotemporal lobar degeneration due to *MAPT* mutation; FTLD TDP-43, frontotemporal lobar degeneration due to TDP-43; FTP, Flortaucipir; LBD, Lewy body dementia; PSP, progressive supranuclear palsy; SUVR, standardized uptake value ratio.

Reviewing individual SUVR images and w-maps (**Supp. Figure 1**), we observed patterns of low-level Flortaucipir binding in non-AD cases either in the white matter or in the basal ganglia, sparing the cortex. All non-AD cases but one LBD case (Braak V, intermediate ADNC levels, included in **Figure 2**) were tau-PET negative based on the visual read performed with FDA-approved guidelines. Quantification of SUVR in the basal ganglia showed higher signal in the substantia nigra, red nucleus, globus pallidum, and subthalamic nucleus in the PSP and CBD groups compared with controls, however with significant overlap between groups (**Supp. Figure 5**).

**Figure 2.**
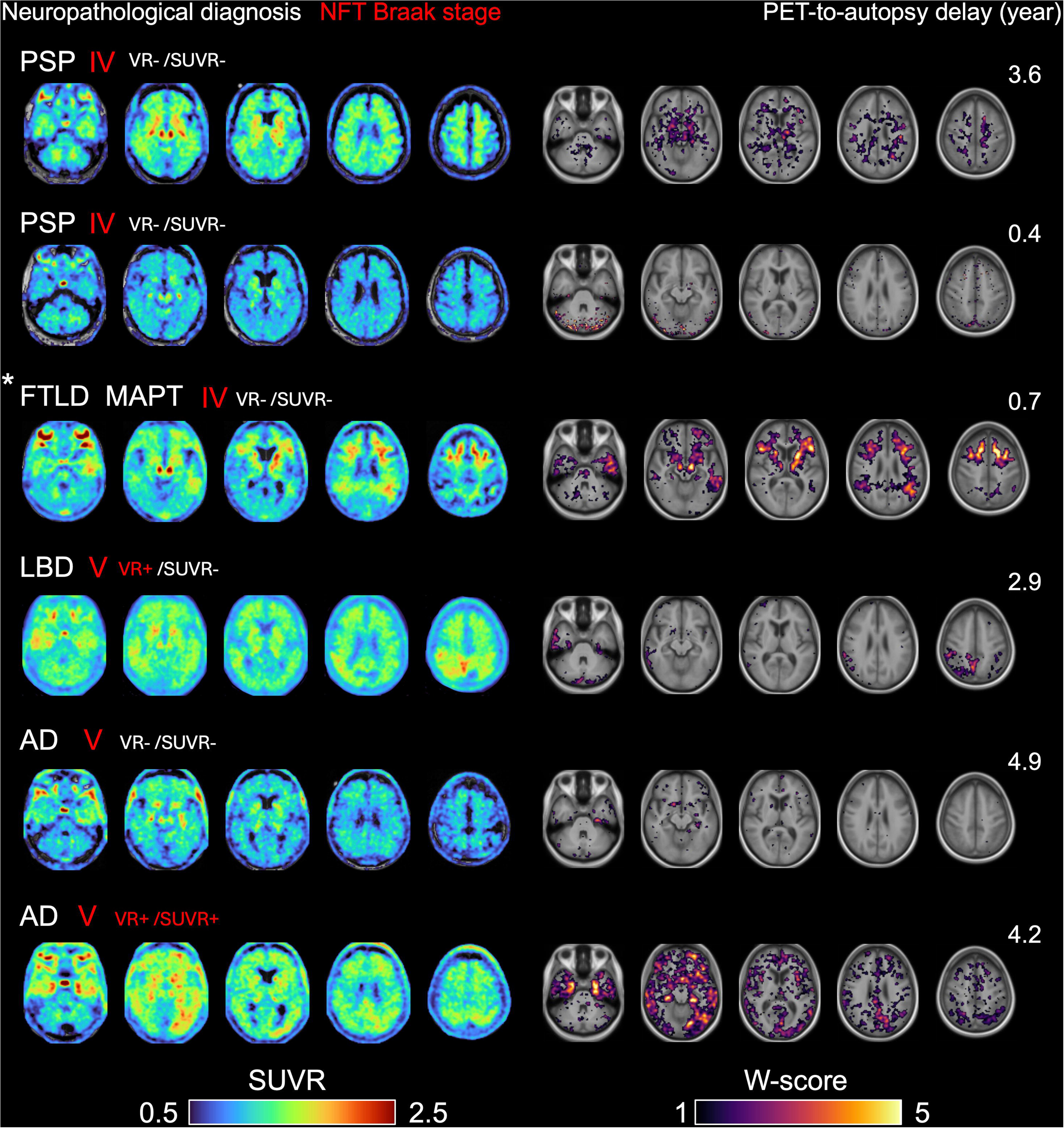
Individual Flortaucipir SUVR images and w-maps for Braak stages III to VI cases. Individual images are presented as axial slices of SUVR images on the left column and individual w-maps on the right column, with each line being one case. *Cases described in Soleimani-Meigooni *et al.*, Brain 2020. All Braak IV (n=3) and V (n=3) cases of the cohort are included. Visual read (VR) per FDA guidelines and quantification (SUVR) positivity (+) and negativity (-) are indicated for each case. W-maps are voxel-wise maps comparing single-subject images to control individuals, adjusting for age and sex (La Joie *et al.*, Journal of Neurosciences 2012). Abbreviations: CBD, corticobasal degeneration; FTLD-*MAPT*, frontotemporal lobar degeneration due to *MAPT* mutation; FTP, Flortaucipir; LBD, Lewy body dementia; NFT, neurofibrillary tangles; PSP, progressive supranuclear palsy; SUVR, standardized uptake value ratio.

### Flortaucipir SUVR association with neuropathology scores

We assessed the association between Flortaucipir SUVRs and NFT Braak stages, Thal phases, and ADNC levels (**Figure 3**).

**Figure 3.**
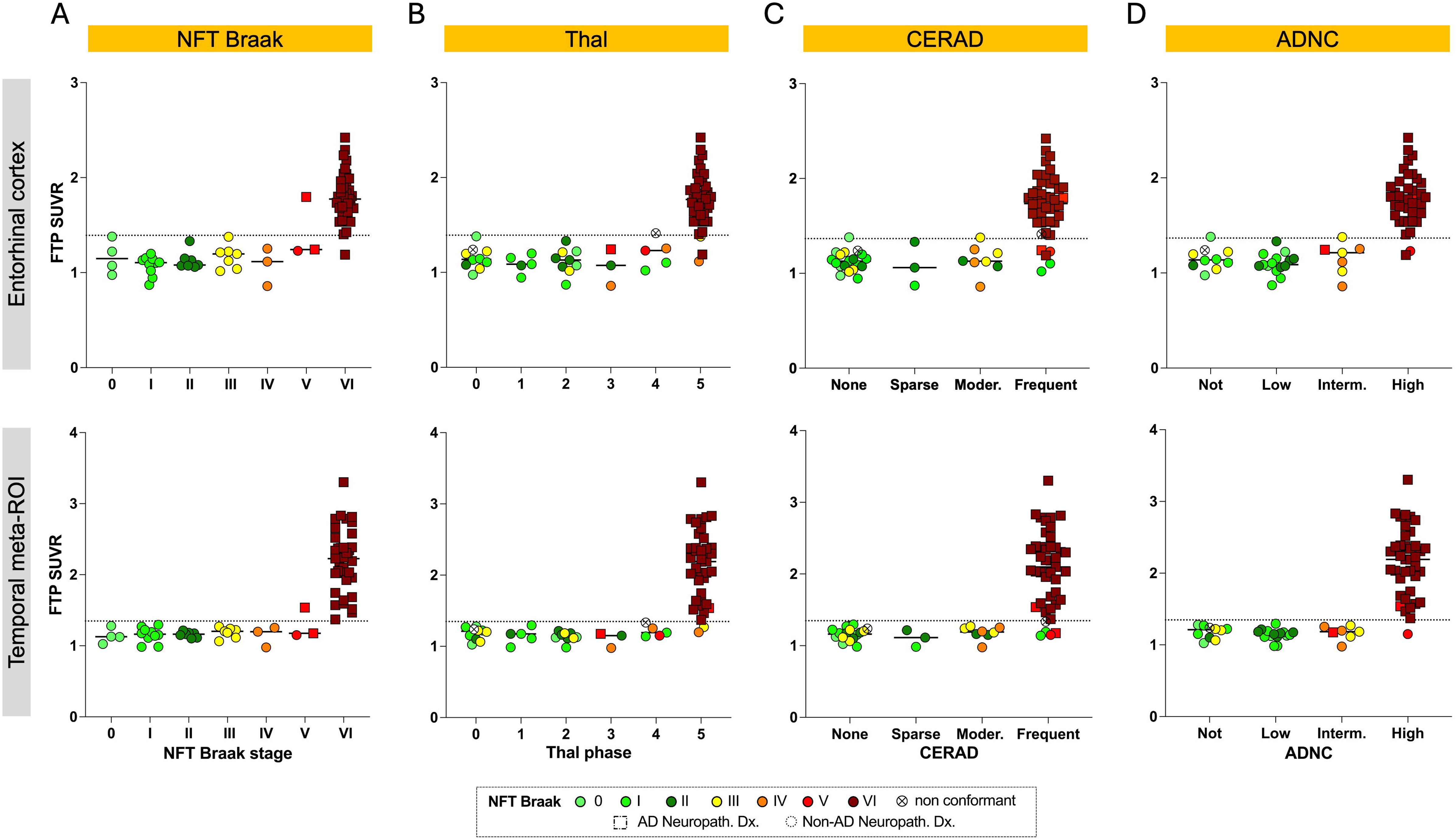
Association of Flortaucipir SUVRs with AD neuropathological changes. Entorhinal cortex SUVR (top row) and temporal meta-ROI SUVR (bottom row) by **A,** NFT Braak stages; **B,** Thal phases; **C,** CERAD score and **D,** levels of ADNC. The horizontal line represents the cut-off for SUVR positivity derived from the control group (mean + 2SD). The n=2 CTE cases were NFT Braak non-conformant, and ADNC levels could not be determined for one of these cases. The Thal phase could not be determined for n=1 CBD case, and subsequently, ADNC levels could not be established. Abbreviations: ADNC, AD neuropathologic changes; FTP, Flortaucipir; NFT, neurofibrillary tangles; SUVR, standardized uptake value ratio.

The NFT Braak stage VI was associated with higher SUVRs in both ROIs compared with controls, above the threshold of positivity (**Figure 3A**). All lower NFT Braak stages (0-IV) displayed SUVRs below the threshold of positivity. All cases with Braak IV (n=3) were below the positivity thresholds for both ROIs, and only one (out of 3) case with Braak V had entorhinal and temporal SUVR values above the threshold. No significant correlations were found between Braak stages and Flortaucipir SUVRs after excluding cases with Braak stage VI (r=0.19-0.26, P≥0.13). All the Braak stages IV and V scans are presented in **Figure 2**. The quantitatively positive Braak V case (primary neuropathology: AD) was visually positive per FDA-approved guidelines for Flortaucipir PET. One Braak V case (primary neuropathology: LBD) was visually positive per FDA reading guidelines due to high uptake (SUVR up to 2.68) in the parietal lobe, including the precuneus, but this was not captured by quantification in the pre-determined temporal ROIs (SUVR= 1.15). All other cases were visually negative.

Regarding association with amyloid pathology, increased Flortaucipir SUVRs were observed in the entorhinal ROI at Thal phase 5 compared with lower Thal phases (**Figure 3B**). Only one non-AD case with Thal 4 (CTE, Braak nonconformant) had elevated Flortaucipir SUVRs in the entorhinal cortex, just above threshold. In the temporal meta-ROI, elevated SUVRs were only associated with Thal phase 5 and only in AD cases. The two non-AD cases with Thal phase 5 displayed SUVRs below the cut-off of positivity. No difference in temporal meta-ROI SUVRs was observed between Thal phases 0 and 4.

For CERAD scores, only AD cases with CERAD frequent displayed elevated Flortaucipir SUVR in our ROIs (**Figure 3C**). No Flortaucipir SUVR elevation was observed in non-AD cases with a CERAD frequent score.

Elevated Flortaucipir SUVRs were associated with high ADNC levels, while low SUVRs were observed in absent, low, and intermediate ADNC levels for both ROIs (**Figure 3D**). The case with high levels of ADNC with negative temporal meta-ROI SUVR had a primary neuropathological diagnosis of LBD. No differences in SUVRs were observed between groups at absent, low, and intermediate ADNC levels. These results were unchanged after applying PVC (**Supp. Figure 4**).

In the non-AD diagnoses, four cases fulfilled the criteria for primary age-related tauopathy (PART; n=3 primary FTLD-TDP cases and n=1 primary LBD case). Additionally, 14 non-AD tauopathy cases had low Thal phase (0-2) with Braak stages ranging from I to IV; however, they did not fulfil PART criteria due to their primary neuropathological diagnosis of FTLD-tau. Flortaucipir SUVRs in the entorhinal ROI and the temporal meta-ROI in both PART and low amyloid/positive Braak cases did not differ from SUVRs in non-autopsy controls and remained below the threshold of positivity (**Supp. Figure 6**).

### Correlation with semi-quantitative tau pathology

Flortaucipir SUVRs were correlated with the semi-quantitative assessment of AD NFT pathology in corresponding brain regions in a subset of 56 participants, including 29 with AD and 27 with non-AD primary neuropathology (**Figure 4**). The global Flortaucipir SUVRs displayed a strong correlation with the average NFT pathology burden (Spearman r=0.82, P<0.001, **Figure 4A**). When considering relationships between semi-quantitative measures of tau lesion and Flortaucipir SUVR in various regions, the same correlations were observed across the different cortical brain regions (Spearman r=0.42-0.86, P ≤0.0016, **Figure 4B, Supp. 7** for detailed plots).

**Figure 4.**
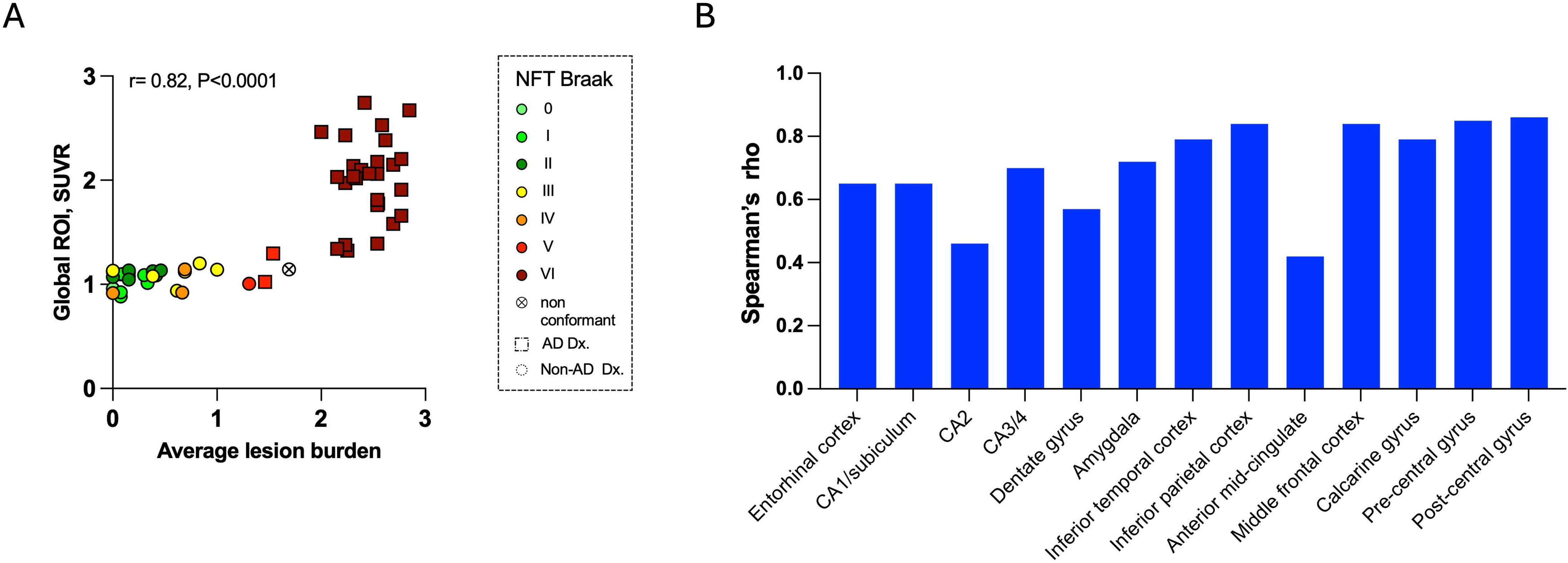
Association of Flortaucipir SUVRs and AD NFT burden. **A**, Scatter plot displaying the association of the global tau SUVR with AD NFT burden. Correlations were analyzed with Spearman’s rank correlations (r). Individual points are color-coded by NFT Braak stages. The global tau ROI corresponds to a large ROI encompassing all Braak stage regions. The average AD NFT burden corresponds to the average burden across 13 cortical brain regions. **B,** Bar chart displaying Spearman’s rank correlation coefficients between FTP SUVRs and AD NFT burden within the 13 cortical brain regions. Anterior midcingulate: P=0.016, CA2: P=0.0004. Other regions: P<0.0001. Abbreviations: FTP, Flortaucipir; NFT, neurofibrillary tangles; SUVR, standardized uptake value ratio.

### Plasma p-tau217 across diagnoses and AD neuropathology scores

Plasma p-tau217 concentrations were measured in 56 cases. The subsample was comparable to the overall sample with tau-PET (**Supp. Table 1**). Flortaucipir SUVRs by diagnoses and the association of Flortaucipir-PET with AD pathology scores in the plasma sample were similar to what was observed in the whole cohort (**Supp. Figure 8**). Plasma p-tau217 concentrations were increased in primary AD cases and LBD cases, compared with other primary non-AD diagnoses (**Figure 5A**). There were no obvious differences in plasma p-tau217 concentrations between the different FTLD diseases.

**Figure 5.**
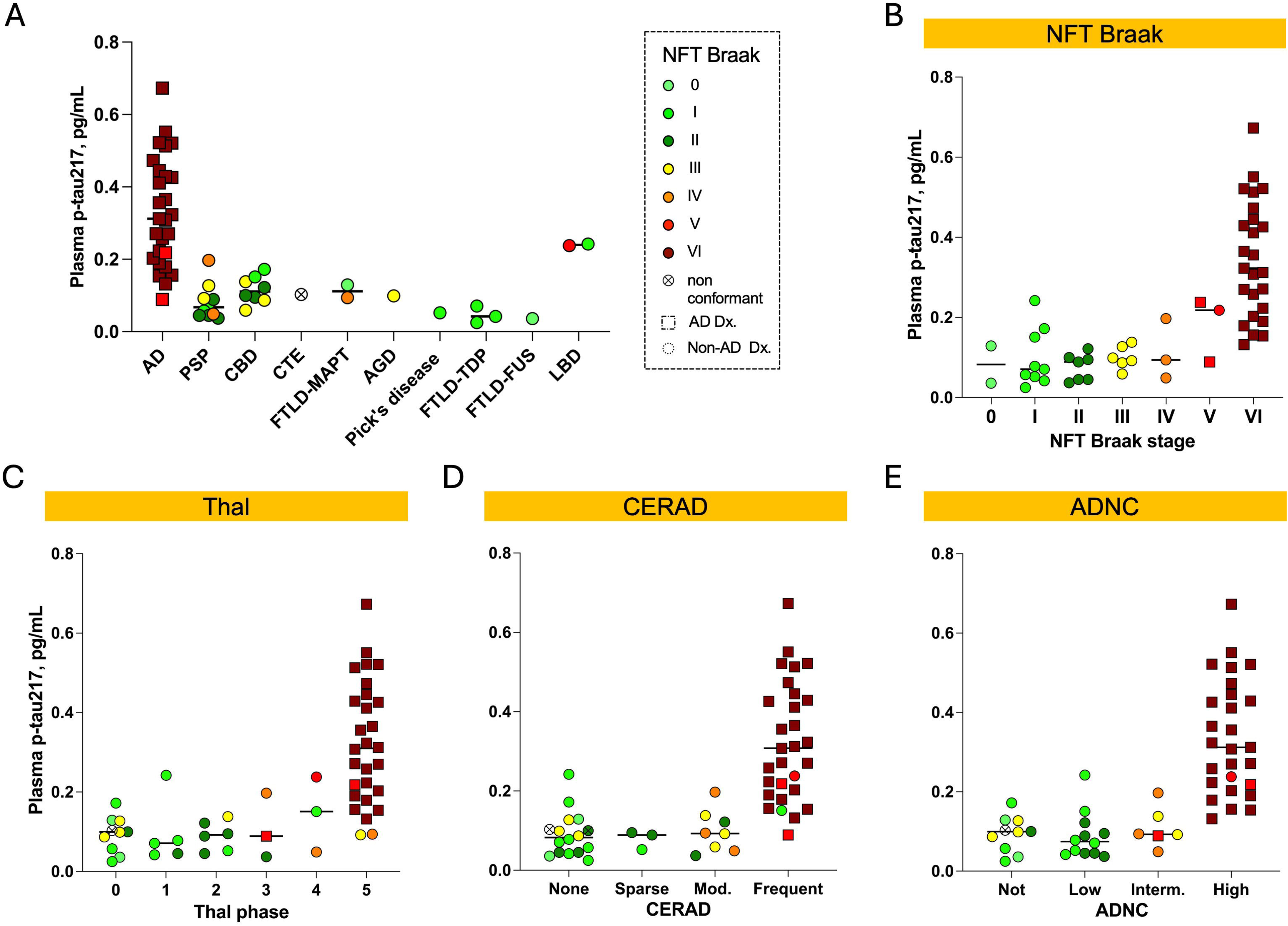
Plasma p-tau217 concentrations by diagnosis and by AD neuropathological scores. **A,** Plasma p-tau217 concentrations by neuropathological diagnosis. Plasma p-tau217 concentrations by **B,** NFT Braak stages; **C,** Thal phases; **D,** CERAD score and **E,** ADNC levels. Abbreviations: AD, Alzheimer’s disease; AGD, argyrophilic grain disease; CBD, corticobasal degeneration; CERAD, Consortium to Establish a Registry for Alzheimer’s Disease; CTE, chronic traumatic encephalopathy; FTLD-*FUS*, frontotemporal lobar degeneration due to a *FUS* mutation; FTLD-*MAPT*, frontotemporal lobar degeneration due to *MAPT* mutation; FTLD-TDP-43, frontotemporal lobar degeneration due to TDP-43; FTP, Flortaucipir; LBD, Lewy body dementia; NFT, neurofibrillary tangles; PSP, progressive supranuclear palsy.

We examined the association of plasma p-tau217 with AD neuropathological scales (**Figure 5B-E**). Higher concentrations of plasma p-tau217 were observed at Braak stage VI compared to all earlier Braak stages (**Figure 5B**). Marginal increases were noted for individual cases at early Braak stages, such as Braak I. Regarding amyloid pathology, plasma p-tau217 concentrations were increased at Thal phase 5 compared to Thal phases 0 to 3 (**Figure 5C**). Two out of three cases at Thal phase 4 displayed plasma p-tau217 concentrations in the same range as those observed at Thal 5. CERAD score frequent was associated with higher concentrations of plasma p-tau217 than CERAD not, low and intermediate scores (**Figure 5D**). Some levels of increase were observed for individual non-AD cases with CERAD score none and moderate.

Plasma p-tau217 concentrations were strongly elevated at high ADNC levels compared with no, low, and intermediate levels (**Figure 5E**). There were no notable differences in plasma p-tau217 concentrations between no, low, and intermediate levels of ADNC.

Looking at the association of the amount of tau pathology (n=51) measured with p-tau217 and semi-quantitative measures of tau, plasma p-tau217 concentrations displayed a significant positive correlation with NFT burden (r=0.75, P<0.0001, **Supp. Figure 9**).

### Head-to-head comparison of plasma p-tau217 and Flortaucipir SUVRs in predicting AD neuropathology

Plasma p-tau217 concentrations displayed a significant association with Flortaucipir SUVRs in both the entorhinal ROI (r=0.75, P<0.001) and the temporal meta-ROI (r=0.77, P<0.001), as displayed in **Figure 6A**. This correlation was sustained by focusing on the AD group in the temporal meta-ROI (r=0.58, P=0.0015). We performed ROC analyses to explore the performance of Flortaucipir SUVRs and plasma p-tau217 concentrations to identify AD neuropathology (**Figure 6B**). Flortaucipir SUVR in both PET ROIs detected advanced Braak stages (V/VI) with a near-perfect performance (both AUC=0.99). Plasma p-tau217 concentrations also displayed high performance with an AUC of 0.96, which did not significantly differ from PET (DeLong test, P=0.15-0.17 compared with PET ROIs). To identify intermediate/high levels of ADNC, the entorhinal ROI, temporal meta-ROI, and plasma p-tau217 concentrations yielded high AUCs (0.99, 0.97, and 0.92, respectively), with no statistical difference between AUCs (DeLong tests, overall P>0.05).

**Figure 6.**
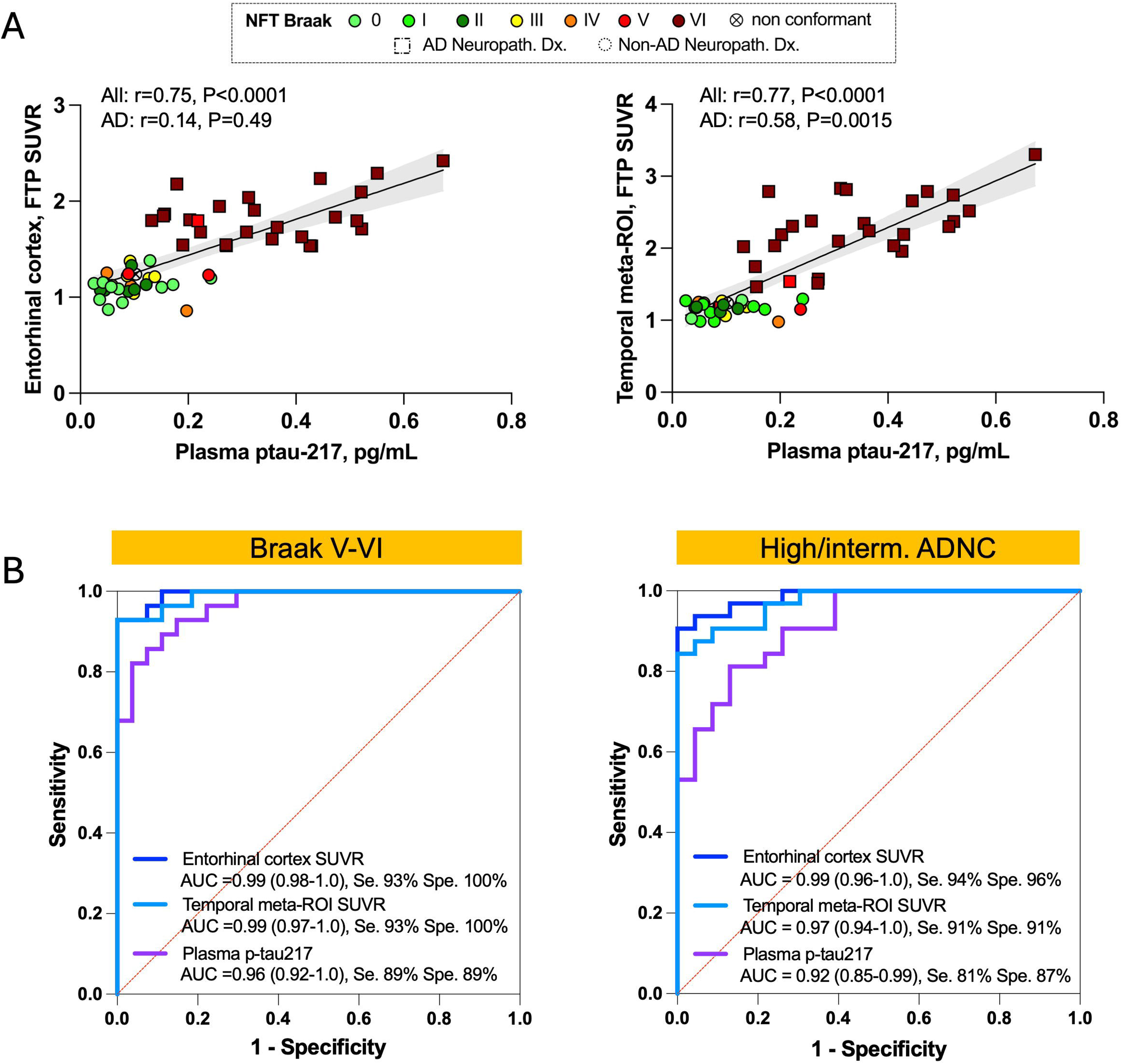
Association of plasma p-tau217 concentrations with Flortaucipir SUVRs and performance to identify AD pathology. **A,** Correlation between plasma p-tau217 concentrations and FTP SUVR ROIs. Correlations were studied using Spearman’s rank correlation (r) in the whole cohort and in the AD subgroup. **B,** ROC analysis of FTP SUVR and plasma p-tau217 performance to identify advanced Braak stages (V/VI) and intermediate/high levels of ADNC. Age, sex, and plasma/PET-to-autopsy delay were added as covariates. Comparison of AUC using the DeLong test: p>0.05. Abbreviations: ADNC, Alzheimer’s disease neuropathologic change; AUC, area under the curve; FTP, Flortaucipir; Se., sensitivity; Spe., specificity; SUVR, standardized uptake value ratio.

## DISCUSSION

This study investigated the comparative diagnostic accuracy of Flortaucipir-PET and plasma p-tau217 concentrations in an autopsy cohort from a tertiary dementia center encompassing a wide range of neurodegenerative diseases with and without tau neuropathology. We found that elevated Flortaucipir-PET signal and high plasma p-tau217 concentrations were associated with advanced Braak stages and high levels of ADNC. There was a significant association of Flortaucipir uptake with AD NFT burden across cortical brain regions. Plasma p-tau217 and Flortaucipir-PET were closely associated and had similar performance in identifying AD pathology. These findings advance our understanding of the association of Flortaucipir-PET and plasma p-tau217 with AD neuropathologic changes.

Here, we confirmed the strong specificity and sensitivity of Flortaucipir-PET for the detection of clinically relevant levels of AD neuropathology in clinically impaired patients evaluated for dementia etiology. Cases with high ADNC levels displayed significantly higher tracer uptake compared with cases with low/intermediate ADNC, detected visually and through quantification.

Patients with non-AD dementia displayed some tracer retention in white matter and/or basal ganglia, with higher tracer uptake than observed in age-matched controls. However, there was no significant elevation in either the entorhinal cortex or the temporal meta-ROI. This aligns with prior studies showing poor correlation between Flortaucipir and non-AD tauopathies, whether 3R or 4R^5–7^. If a majority of these had some level of comorbid amyloid pathology and/or NFTs, amyloid lesions were mostly non-neuritic, and NFT Braak stages were ≤ IV. One LBD case displayed high ADNC, which is a common co-pathology -- their Flortaucipir-PET scan showed parietal-predominant tracer uptake, which was visually positive following the FDA-approved method, but was not captured by quantification in the two temporal ROIs^35^. Similarly, cases with PART did not show elevated Flortaucipir signal. This finding that Flortaucipir does not detect Braak stages I to III is concordant with previously published studies^2,4,5,36^. Findings at Braak IV and V stages vary. An elevated Flortaucipir signal may be found in the entorhinal cortex starting at Braak IV^2,4,37^. In a large autopsy cohort, Josephs *et al*. observed that amyloid pathology levels mediated Flortaucipir uptake at all Braak stages in the entorhinal cortex: Flortaucipir-PET signal only increased at intermediate Braak stages in the presence of a high Thal phase^37^. In our cohort, cases with intermediate Braak displayed low levels of amyloid pathology, which could have contributed to the low Flortaucipir signal in those cases. It is also possible that the NFT density was lower in our Braak IV-V cases with non-AD diagnosis compared with patients with pure AD pathology at equal Braak stage. The NFT Braak staging system is purely topographical, and studies have shown substantial variations in the amount of pathology for a given Braak stage^38^. We also cannot exclude that some non-AD 3R- or 4R-tau lesions and associated neurodegenerative processes may have affected tracer binding to AD-tau neuropathology.

Using a semi-quantitative visual assessment, Flortaucipir-PET uptake was correlated with AD NFT burden in a non-linear fashion. Increased tracer uptake across cortical regions was associated with a higher AD-tau lesion burden that was only observed in AD cases corresponding to Braak stage VI. This is in line with prior autoradiography studies that have found strong and diffuse binding to NFTs in high Braak stage samples^39,40^. We observed some levels of NFT in the non-AD cases; however, they were most likely not sufficient for Flortaucipir-PET detection, especially in the FTLD cases. Indeed, some authors have established region-dependent thresholds of tau burden for Flortaucipir-PET tracer uptake^38^. Thus, Flortaucipir appears to lack sensitivity to detect mild levels of AD copathology associated with low tau burden.

A large number of studies have explored the association of plasma p-tau217 with tau PET^9,12,13^. However, there is limited data relating both biomarkers to autopsy findings.

In the whole sample, we observed significant correlations between plasma p-tau217 levels and Flortaucipir SUVRs, which is largely consistent with the existing literature. However, correlations were weaker when focusing on the subgroup with AD, and the correlation was only significant between plasma p-tau217 concentrations and Flortaucipir SUVRs in the temporal meta-ROI, not in the entorhinal ROI. Indeed, studies have reported a significant variability in the global tau PET burden for a given p-tau217 concentration at the individual levels^12,14^. One aspect is most likely that plasma p-tau species not only reflect neurofibrillary tangles but also amyloid pathology, especially early in the AD continuum^8,11^. Thus, plasma p-tau217 and tau PET, even if highly associated, are not identical in their relationship with tau burden.

Plasma p-tau217 concentrations were strongly associated with AD pathology. We observed consistently elevated p-tau217 concentrations at advanced Braak stages and high levels of ADNC, in a similar profile to PET, with a significant correlation with NFT burden^10,11,41,42^. Previous studies reported that plasma p-tau becomes abnormal before tau-PET^43^. Flortaucipir-PET and plasma p-tau markers might relate to different AD neuropathologic features, which may explain this phenomenon. Prior studies have shown that amyloid pathology could trigger increased production, phosphorylation, and release of soluble phosphorylated tau, which occurs before the presence of a tau PET signal^44,45^. However, our results did not suggest a significantly higher sensitivity of plasma p-tau217 for intermediate AD-tau pathology. Prior works did observe some elevation of plasma p-tau217 at low/intermediate ADNC levels, albeit with a significant overlap with concentrations observed in the absence of ADNC^10,11^. Different p-tau species have been reported to have specific trajectories of changes in earlier stages of AD pathology, such as plasma p-tau231, which could have better detected lower ADNC levels^46,47^. Novel biomarkers such as plasma MTBR-tau243, thought to specifically reflect tau tangle pathology, may also offer better estimates of the tau burden in AD^48^.

Overall, our data supports that plasma p-tau217 can perform as an effective and accessible screening alternative to neuroimaging to identify participants with advanced AD pathology, especially in places with limited access to PET imaging, due to similar performance compared to tau PET, and its relatively lower cost. However, tau PET still provides unique spatial information on tau distribution as well as a validated proxy for evaluation of tau burden.

### Limitations

This work has several limitations. Our cohort originated from a tertiary dementia center with recruitment targeting young-onset dementia, with a high prevalence of FTLD diagnoses. The presence of non-AD proteinopathies and significant neurodegeneration may have affected the threshold for tracer detection. Thus, our findings may not directly translate to patients in the prodromal phase of AD or to older patients. Cohorts containing larger numbers of cases with intermediate NFT Braak stages and with limited co-pathologies will be needed to give more definite answers on Flortaucipir-PET sensitivity. We should also consider the limitations of the NFT Braak staging. First, some studies have identified AD and non-AD cases with alternative distributions of NFT. Secondly, NFT Braak staging is a topographical staging system and does not represent a linear quantitation of pathology. Thus, it might not be a sufficient measurement against which to evaluate the sensitivity of tau PET. More continuous measures of neuropathological burden, including new methods based on digital neuropathology, would be needed to better capture the threshold of tau pathology associated with Flortaucipir uptake^49^. As our median PET-to-autopsy delay was over 3 years, the tau burden may have increased by the time of autopsy. Plasma p-tau217 measurements and semi-quantitative measurements of tau pathology were missing for a smaller part of the cohort. However, neither these subsamples nor the whole cohort differed in the range of diagnosis or pathological profiles. Finally, there was no autopsy data for our amyloid-negative controls, for which we cannot exclude very early AD pathology or the presence of other proteinopathies.

## Conclusion

In conclusion, our findings provide additional evidence that Flortaucipir-PET has high specificity for AD neuropathology and good sensitivity for high ADNC. Flortaucipir-PET was associated with measures of AD NFT. Plasma p-tau217 was highly concordant with tau PET to detect advanced ADNC, supporting its use for diagnosis and as an accessible biomarker for participant screening and inclusion for clinical trials targeting AD pathologies. However, plasma p-tau217 displayed only a moderate association with the overall tau burden as measured by Flortaucipir PET. Finally, both Flortaucipir-PET and plasma p-tau217 lacked sensitivity to detect early AD-related tau co-pathology in patients with a primary non-AD neuropathological diagnosis.

## Supporting information

All Supplementary material combined

## Abbreviations

Aβ: amyloid-beta;
ADNC: Alzheimer’s disease neuropathological change;
AGD: argyrophilic grain disease;
AUC: area under the curve;
CBD: corticobasal degeneration;
CTE: chronic traumatic encephalopathy;
CERAD: consortium to establish a registry for Alzheimer’s disease;
FTLD: frontotemporal lobar degeneration;
FTP: [^18^F]Flortaucipir;
FUS: RNA-binding protein fused in sarcoma/translocated in liposarcoma;
LBD: Lewy body dementia;
MMSE: mini-mental state examination;
MAPT: microtubule-associated protein tau;
NFT: neurofibrillary tangle;
PART: primary age-related tauopathy;
PET: Positron emission tomography;
p-tau217: phosphorylated tau at threonine 217;
PSP: progressive supranuclear palsy;
PVC: partial volume correction;
ROC curve: receiver operating characteristic curve;
ROI: region of interest;
SUVR: standardized uptake value ratio;
TDP-43: TAR DNA binding protein 43.

## STATEMENTS

### Author Contributions

**Drafting/revision of the manuscript for content, including medical writing for content**: all authors

**Major role in the acquisition of data:** Salvatore Spina, Tia Lamore, Claire Yballa, David N. Soleimani-Meigooni, Ganna Blazhenets, Adam L. Boxer, Julio Rojas-Martinez, Argentina Lario Lago, William J. Jagust, Bruce Miller, Howard J. Rosen, William W. Seeley, Lea T Grinberg, Gil Dan Rabinovici, Renaud La Joie

**Study concept or design**: Agathe Vrillon, Salvatore Spina, David N. Soleimani-Meigooni, Lea T Grinberg, Gil Dan Rabinovici, Lawren Vandevrede, Renaud La Joie

**Analysis or interpretation of data:** Agathe Vrillon, Salvatore Spina, Jhony Mejía-Pérez, David N. Soleimani-Meigooni, Ganna Blazhenets, Lea T Grinberg, Gil Dan Rabinovici, Lawren Vandevrede, Renaud La Joie

