## Supplementary material for "Association of [^18^F]Flortaucipir-PET and plasma p-tau217 with tau neuropathology in AD and other neurodegenerative disorders": All Supplementary material combined

**SUPPLEMENTAL MATERIAL**

e-Methods

Supplementary Figure 1, Individual F Flortaucipir SUVRs images and W-maps

Supplementary Figure 2, Association of Flortaucipir SUVRs with neuropathological diagnoses and AD pathology score, displaying SUVR cut-offs derived from the control group 95th percentile

Supplementary Figure 3, Flortaucipir SUVRs across Braak ROIs by neuropathological diagnosis

Supplementary Figure 4, Association of neuropathological diagnoses and ADNC scores with Flortaucipir-PET after partial volume correction

Supplementary Figure 5, Flortaucipir SUVRs in basal ganglia ROIs by neuropathological diagnosis

Supplementary Figure 6, Supplementary Figure 6, Flortaucipir SUVRs in PART and low-amyloid/Braak positive FTLD-tau cases

Supplementary Figure 7, Association of the AD NFT burden with Flortaucipir SUVRs across cortical regions

Supplementary Figure 8, Flortaucipir-PET SUVRs across diagnoses and association with pathological scores in the plasma sample

Supplementary Figure 9, Association of plasma p-tau217 with AD NFT burden

Supplementary Table 1, Demographics and plasma biomarker values in the plasma subsample (n=56)

**eMethods**

*Clinical diagnosis*

All participants included underwent at least one in-depth evaluation. Many participants had several visits with cognitive follow-up. Clinical diagnoses were made by consensus application of standard research criteria ^1–7^.

*Genetic assessment*

Patients with AD underwent genetic screening for *PSEN1* and *PSEN2*, amyloid precursor protein (*APP*), microtubule-associated protein tau (*MAPT*), chromosome 9 open reading frame 72 (*C9orf72*), RNA-binding protein fused in sarcoma/translocated in liposarcoma (*FUS)*, and progranulin (*GRN*). Patients in the FTLD spectrum underwent gene screening for *MAPT, C9orf72, FUS,* and *GRN.* All but two participants (71/73) underwent APOE genotyping. Individuals with one or two *APOEε4* alleles were classified as carriers versus noncarriers.

FTLD-MAPT cases included n=1 case due to a 305I mutation, n=1 to a P301L mutation and n=1 to a IVS10+16 4R mutation. These mutations have been associated to underlying 4R tauopathie neuropathology ^8–10^.

FTLD TDP-43 cases included n=2 cases due to a *C9orf72* mutation (respectively TDP-43 type B and type U) and n=1 case due to a *GRN* mutation (TDP-43 type A).

*MRI*

Patients underwent morphological MRI on a 3T scanner, on a Siemens TRIO (n=25) or Siemens Prisma (n=48). T1-weighted magnetization-prepared rapid gradient-echo (MPRAGE) MRI sequences (voxel size=1 mm isotropic; matrix=240 × 256 X 160 mm; repetition time=2300 ms; inversion time = 900 ms; flip angle = 9°; echo time = 2.98 ms for Trio and 1.9 for Prisma) were used for PET preprocessing. In BACS controls, MRI was acquired on a Siemens TRIO (voxel size=1 mm isotropic, matrix=256 x 240 x 176 mm; repetition time=2300 ms; inversion time=900 ms, flip angle=9°; echo time=2.96 ms).

*Basal ganglia ROI*

FTP-SUVRs in basal ganglia ROIs were obtained using WFU PickAtlas^11,12^. FTP-PET images were spatially normalized to the Montreal Neurological Institute template in SPM7. The following ROIs were extracted: substantia nigra, red nucleus, globus pallidum, putamen, caudate, subthalamic nucleus and thalamus and midbrain.

*W-maps*

Voxelwise w-score (i.e. covariate-adjusted Z-score) maps were computed from the FTP-PET SUVR images in standardized MNI space for each case. W-scores compare each patient’s SUVR in a given voxel to the SUVR value expected for the patient’s age and sex, based on the amyloid-negative cognitively unimpaired control group^13^.

*Amyloid PET*

N=66 participants received an injection of ~15 mCi [11C] Pittsburgh compound-B (PiB) and images were acquired on the Siemens Biograph PET/CT scanner at Lawrence Berkeley National Laboratory (LBNL). SUVR maps were created from 20-minute PET acquisitions. Cerebellar gray matter was used as the reference region, defined using MRI and Freesurfer.

Three patients received an injection of ~10 mCi [18F]Florbetapir and underwent a 50- to 70-minute post-injection acquisition on the GE Discovery STE/VCT PET/CT scanner at UCSF. The whole cerebellum was used as the reference region^14^.

One patient received an injection of ~8 mCi [18F]Florbetaben and underwent a 90-to-110-minute post-injection acquisition on the GE Discovery STE/VCT PET/CT scanner at UCSF. The whole cerebellum was used as the reference region^14^.

The average SUVR was extracted from a large neocortical region of interest (ROI) encompassing frontal, cingulate, temporal, and parietal areas and subsequently converted into Centiloid values using previously validated and published methods^15,16^. Amyloid-positivity was determined by visual interpreatation by a certified clinician according to FDA–approved criteria for each tracer.

*Supplementary references*

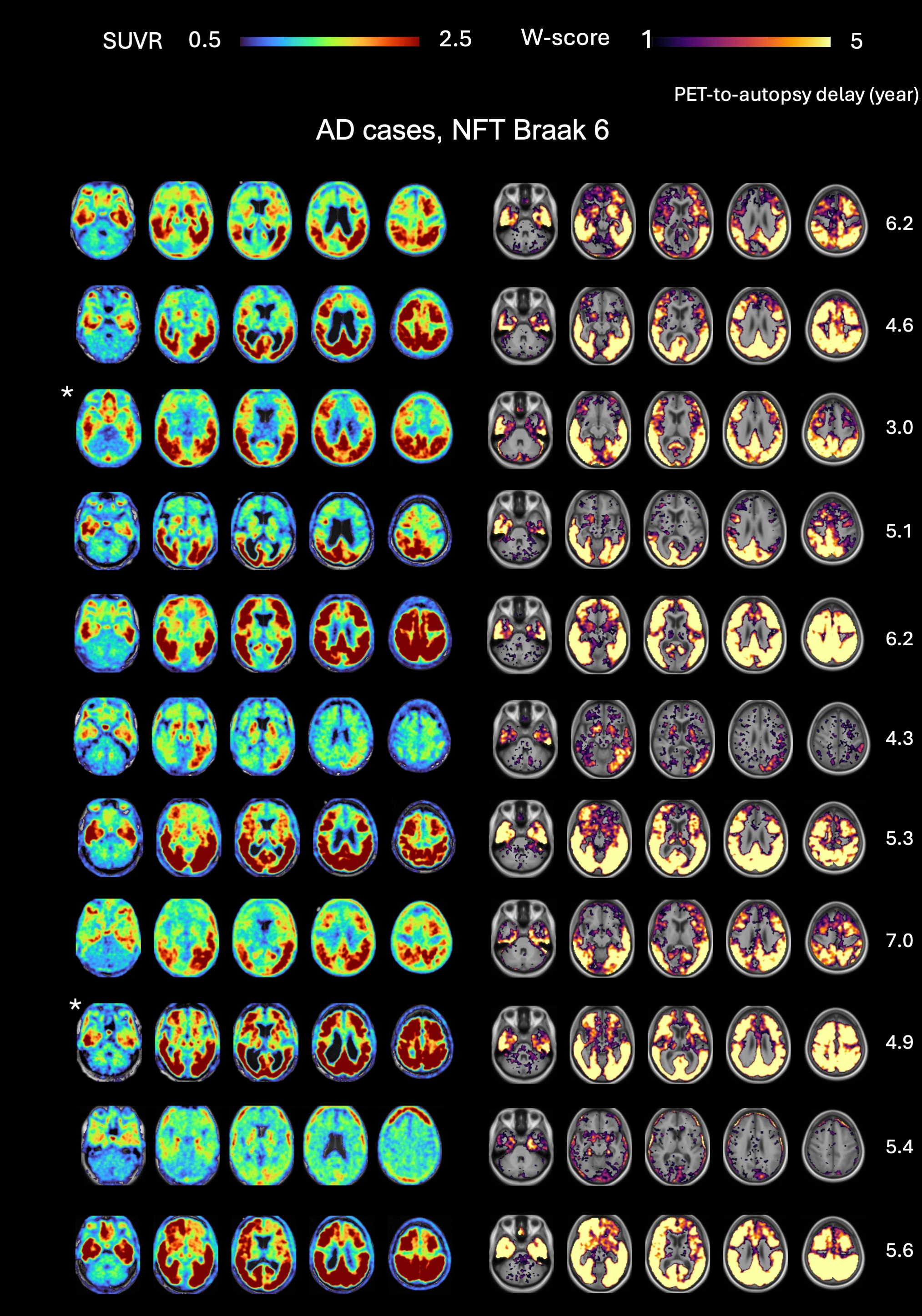

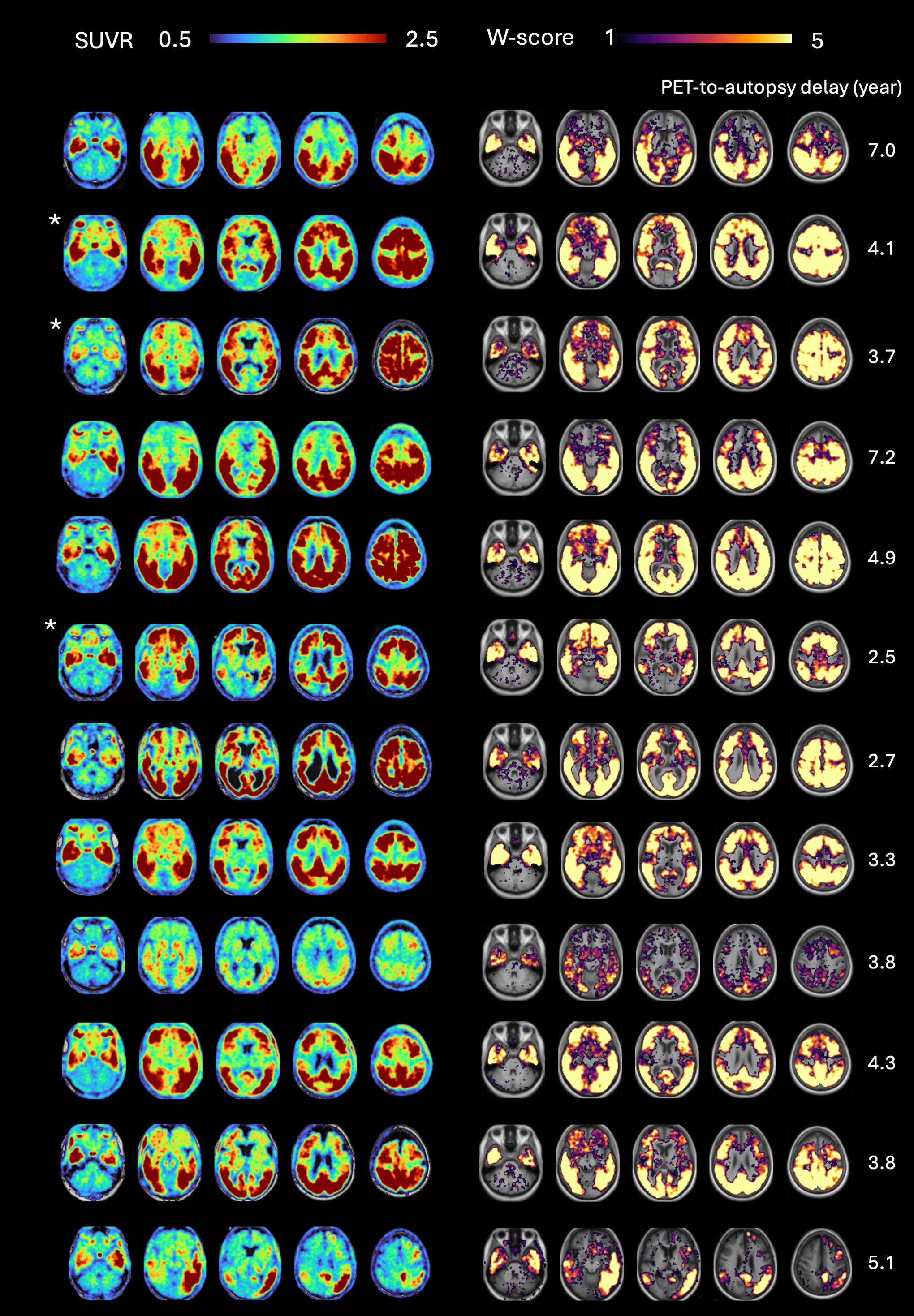

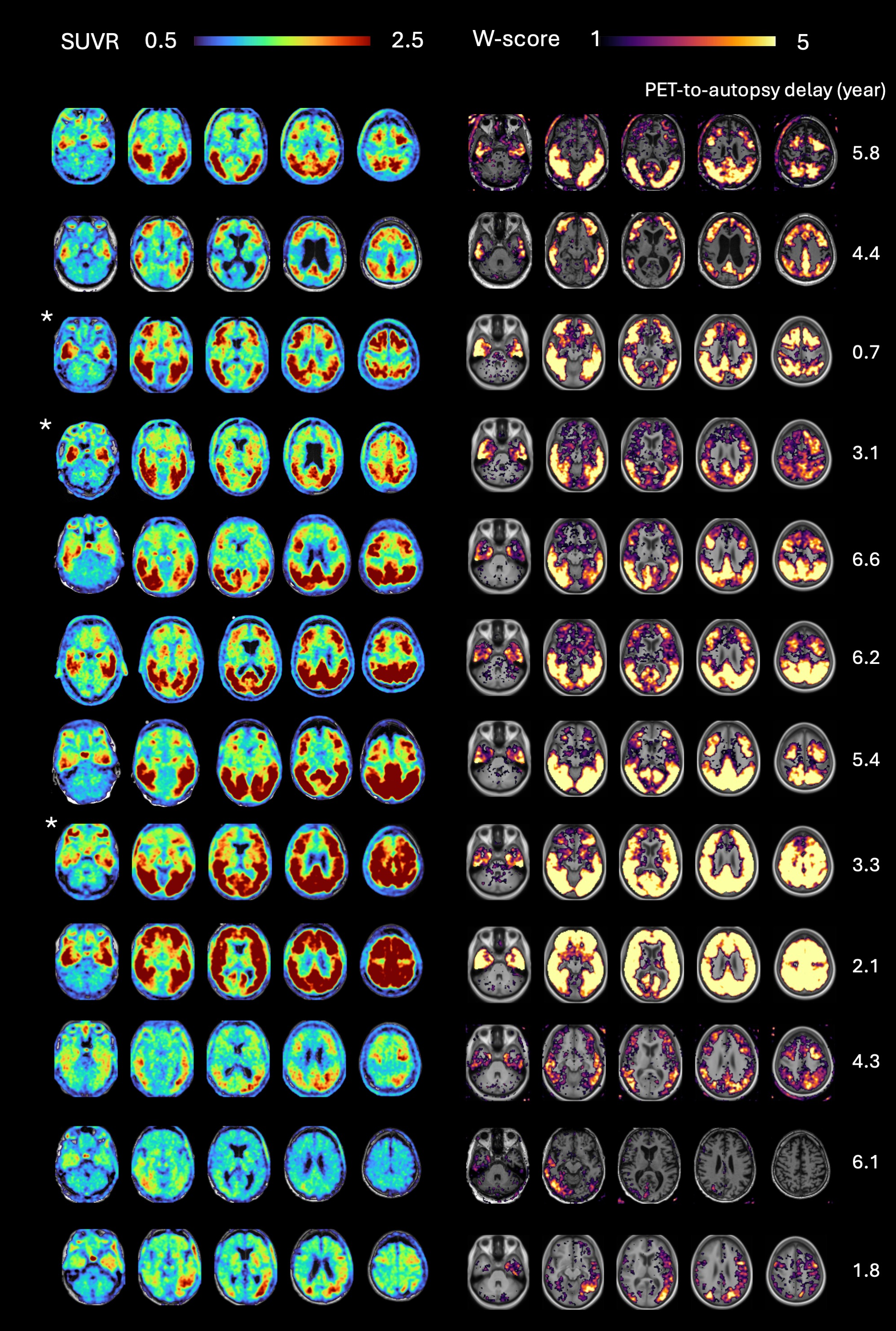

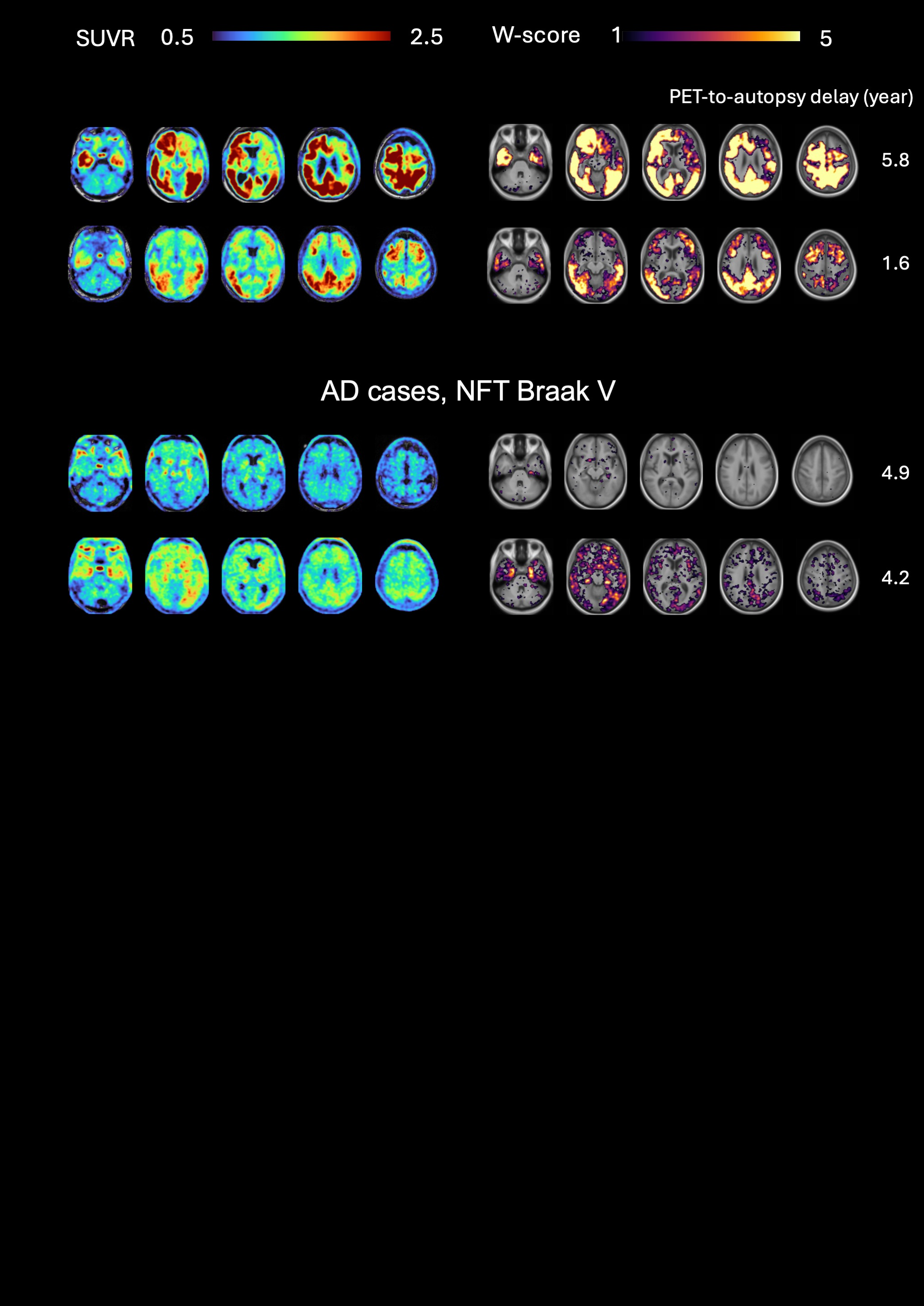

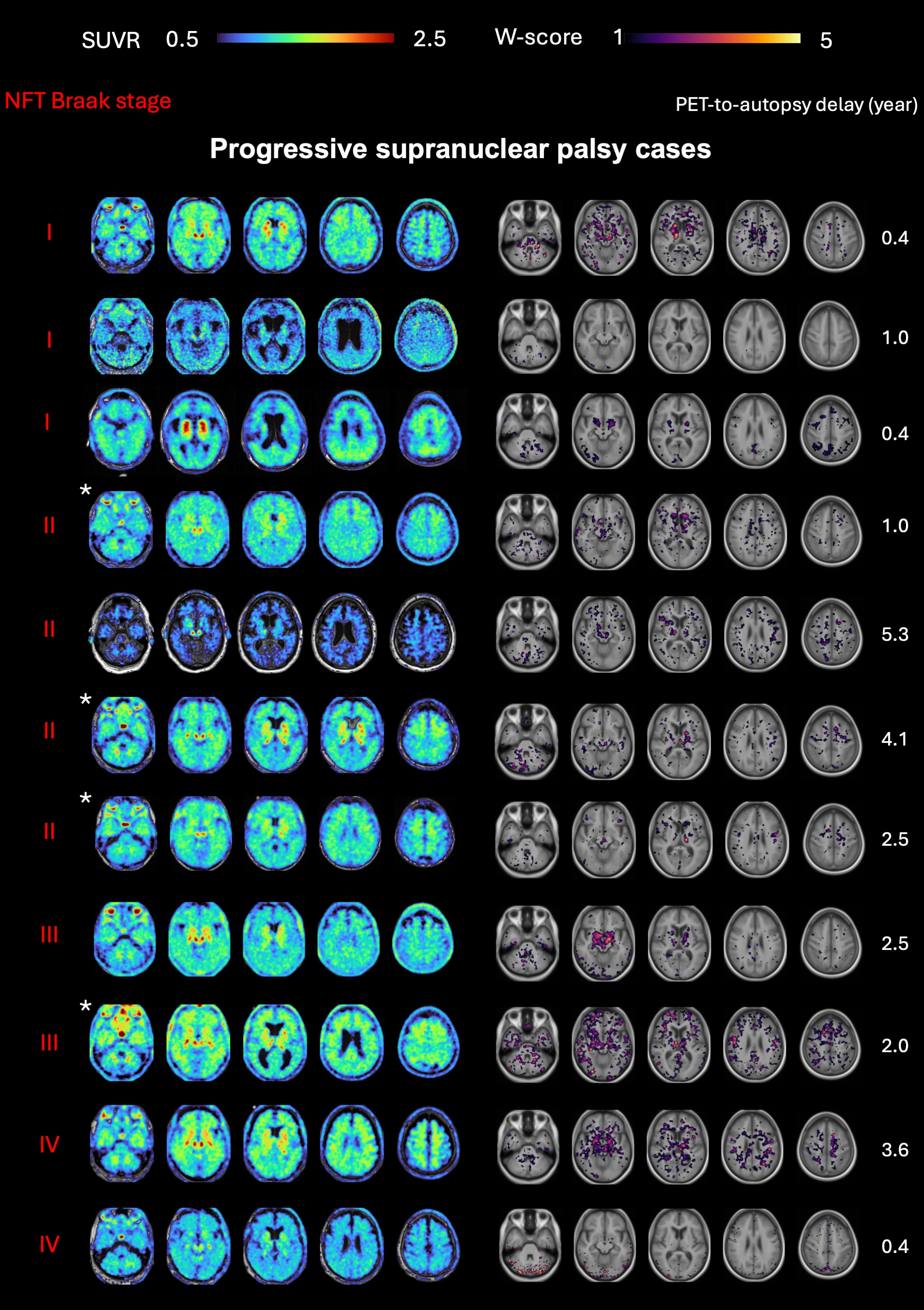

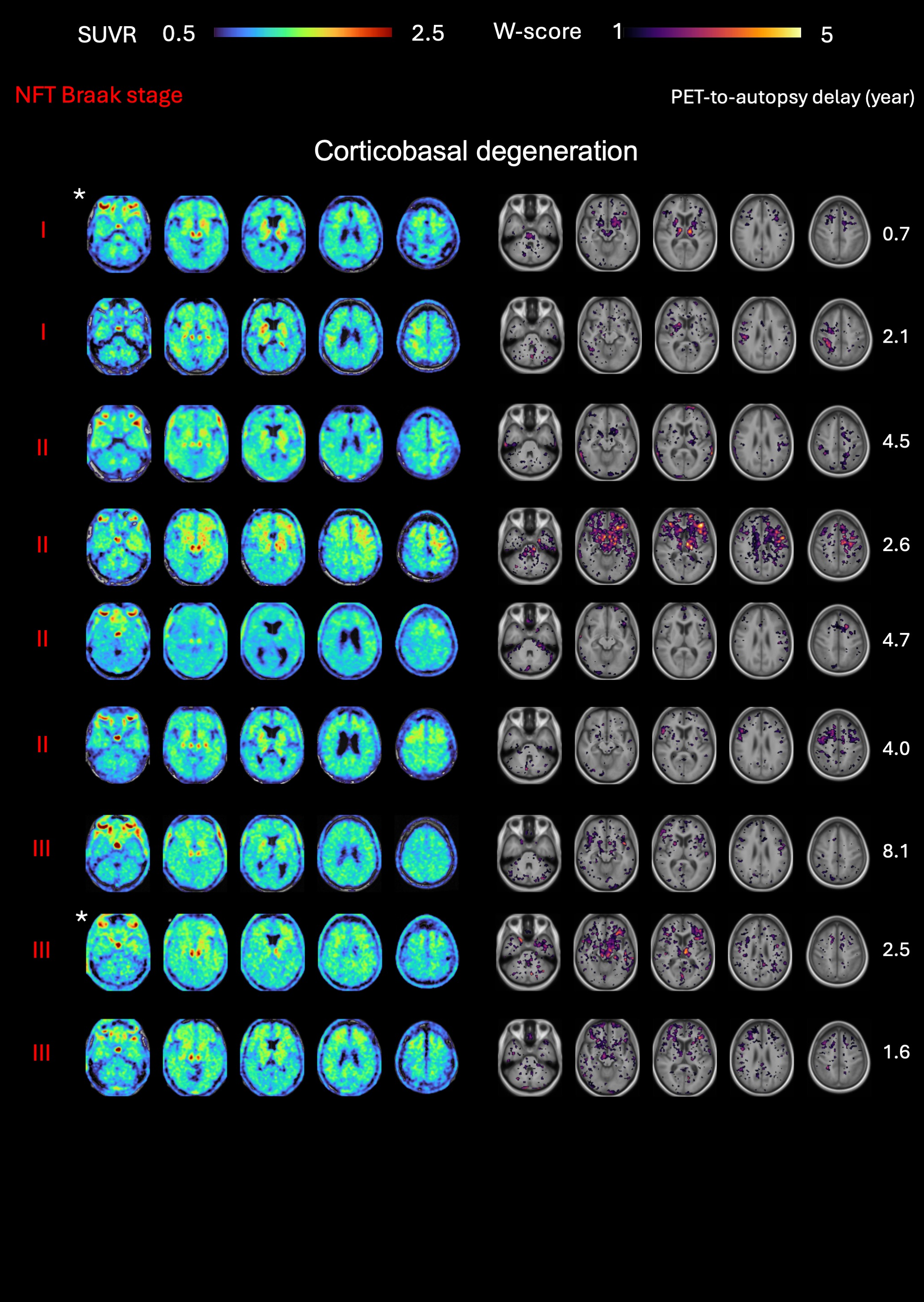

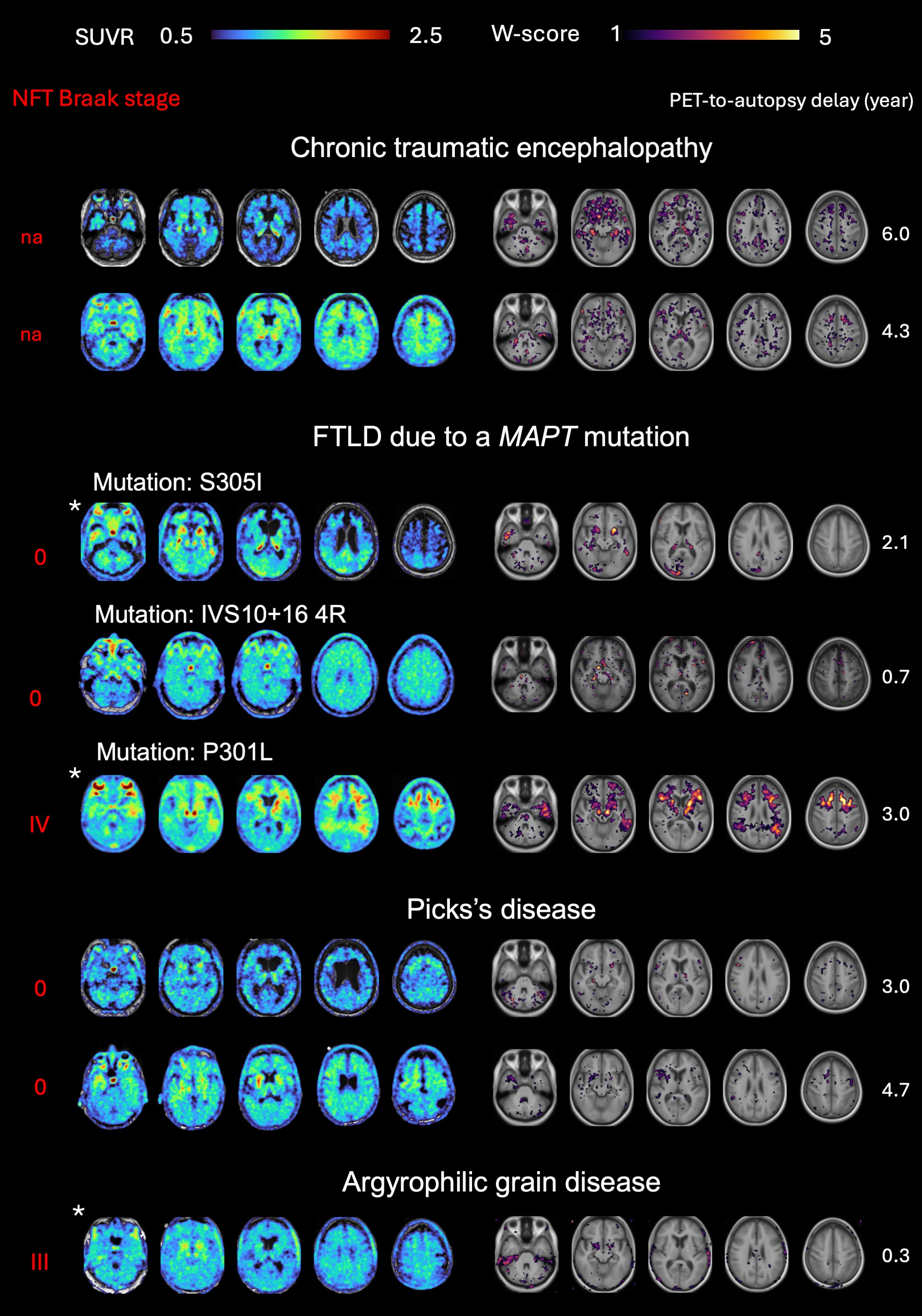

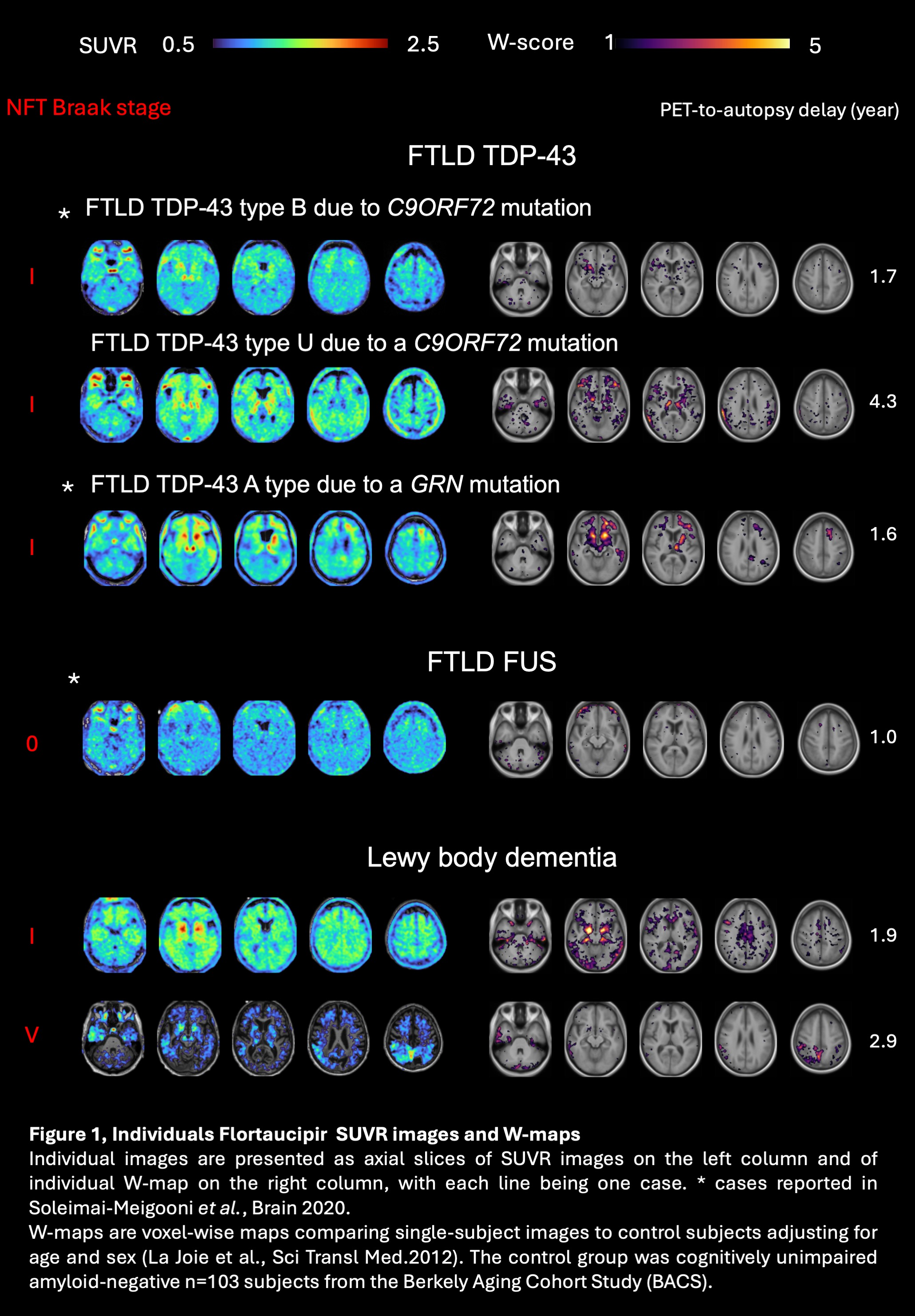

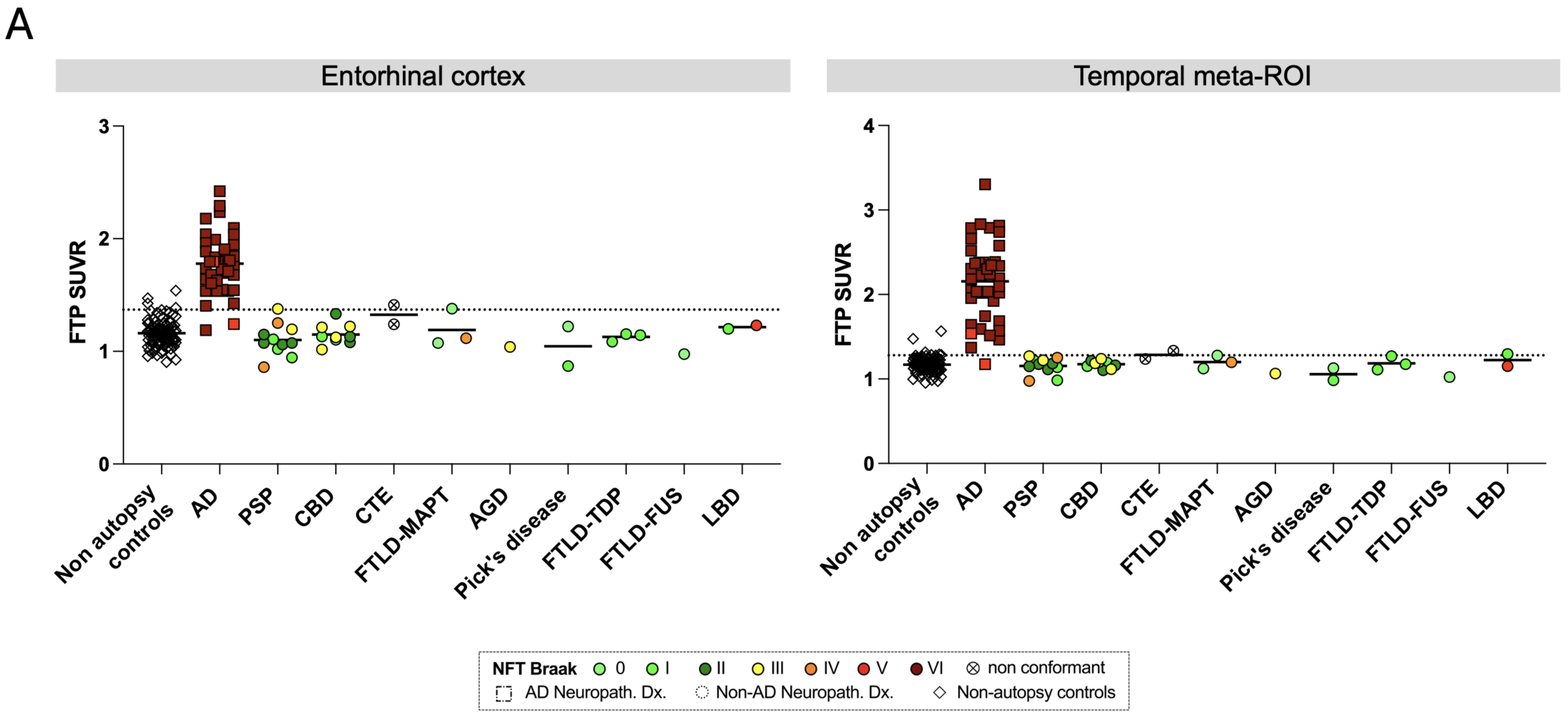

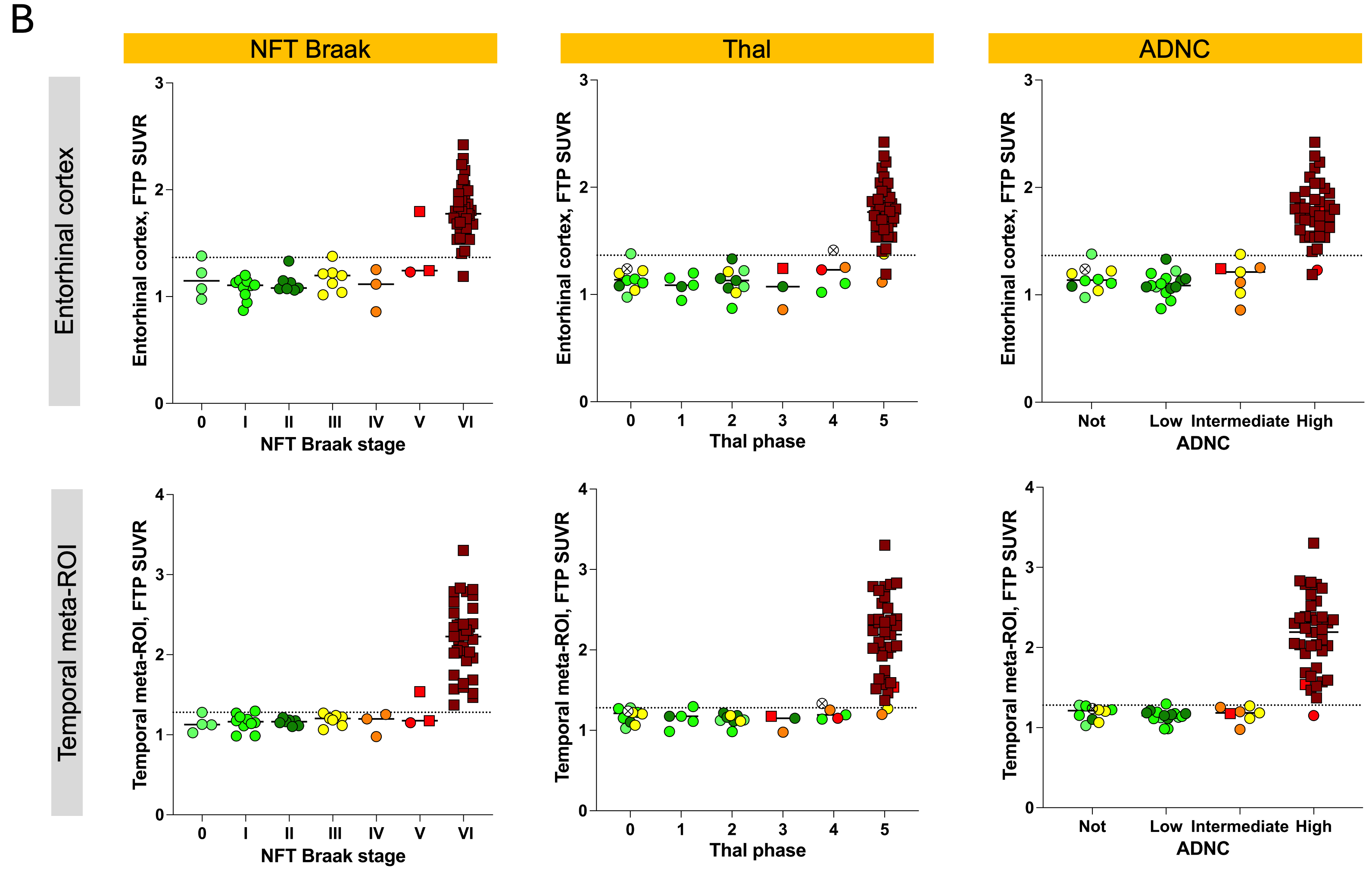

**Supplementary Figure 2, Association of Flortaucipir SUVRs with pathological diagnoses and AD pathology score, displaying SUVR cut-offs derived from the control group 95^th^ percentile (instead of mean + 2SD as in the main paper).**

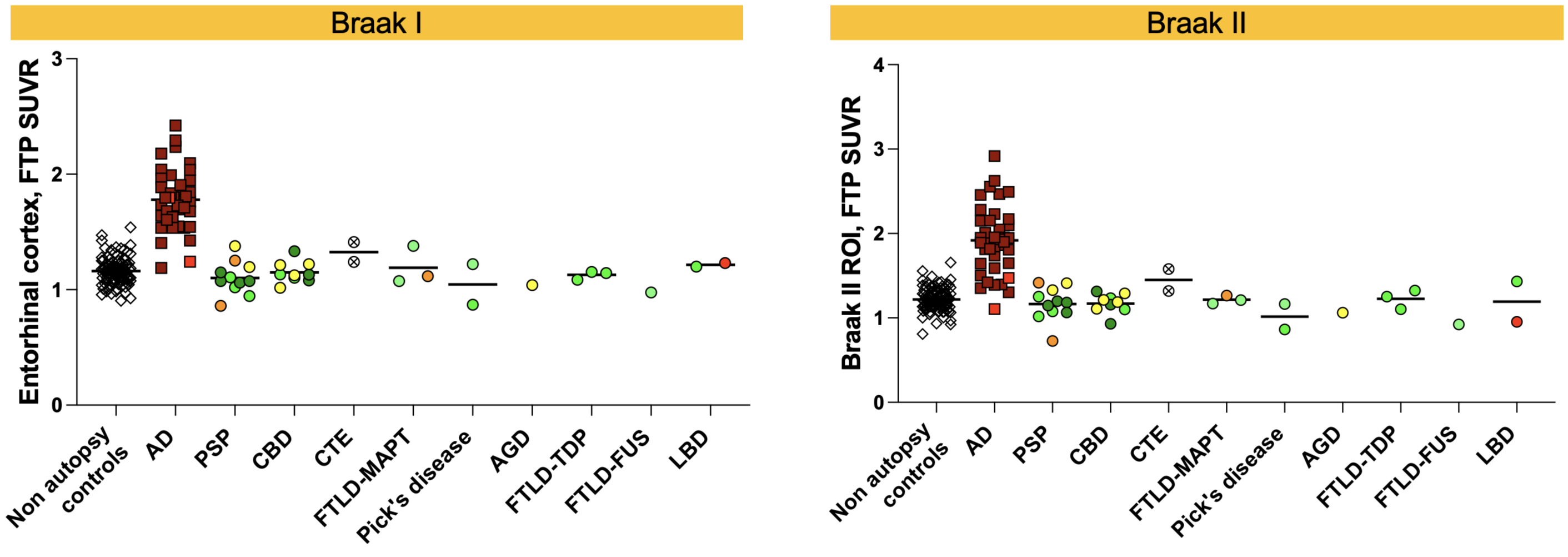

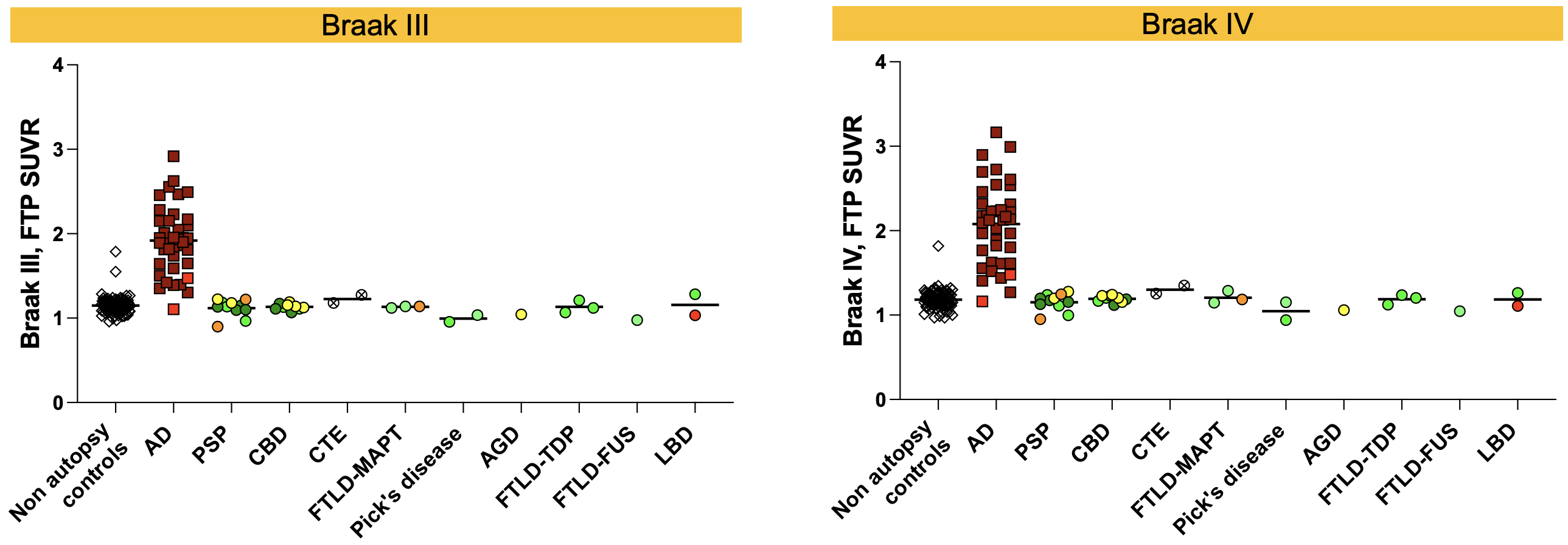

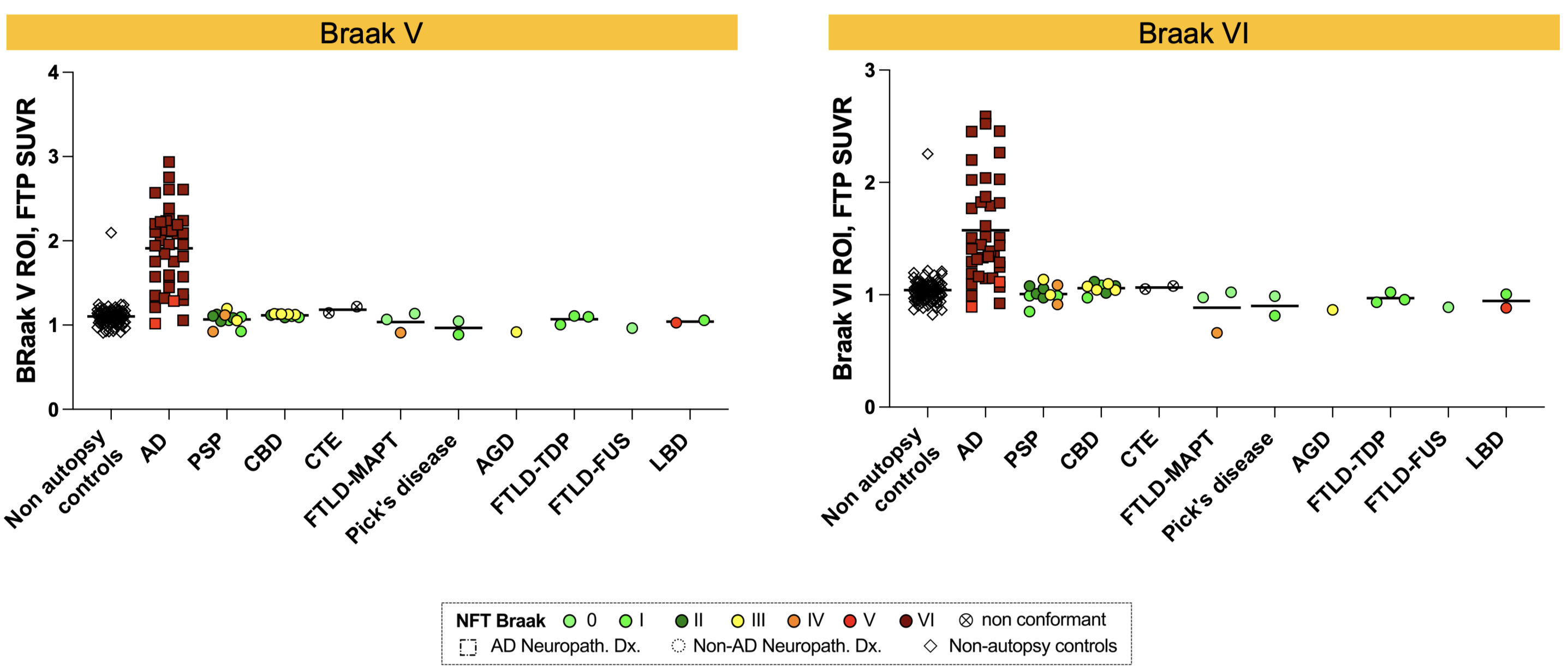

**Supplementary Figure 3, Flortaucipir SUVRs across Braak ROIs by neuropathological diagnosis**

Individual points are color-coded by Braak stages. Rounds indicate non-AD diagnosis, and squares indicate AD diagnosis.

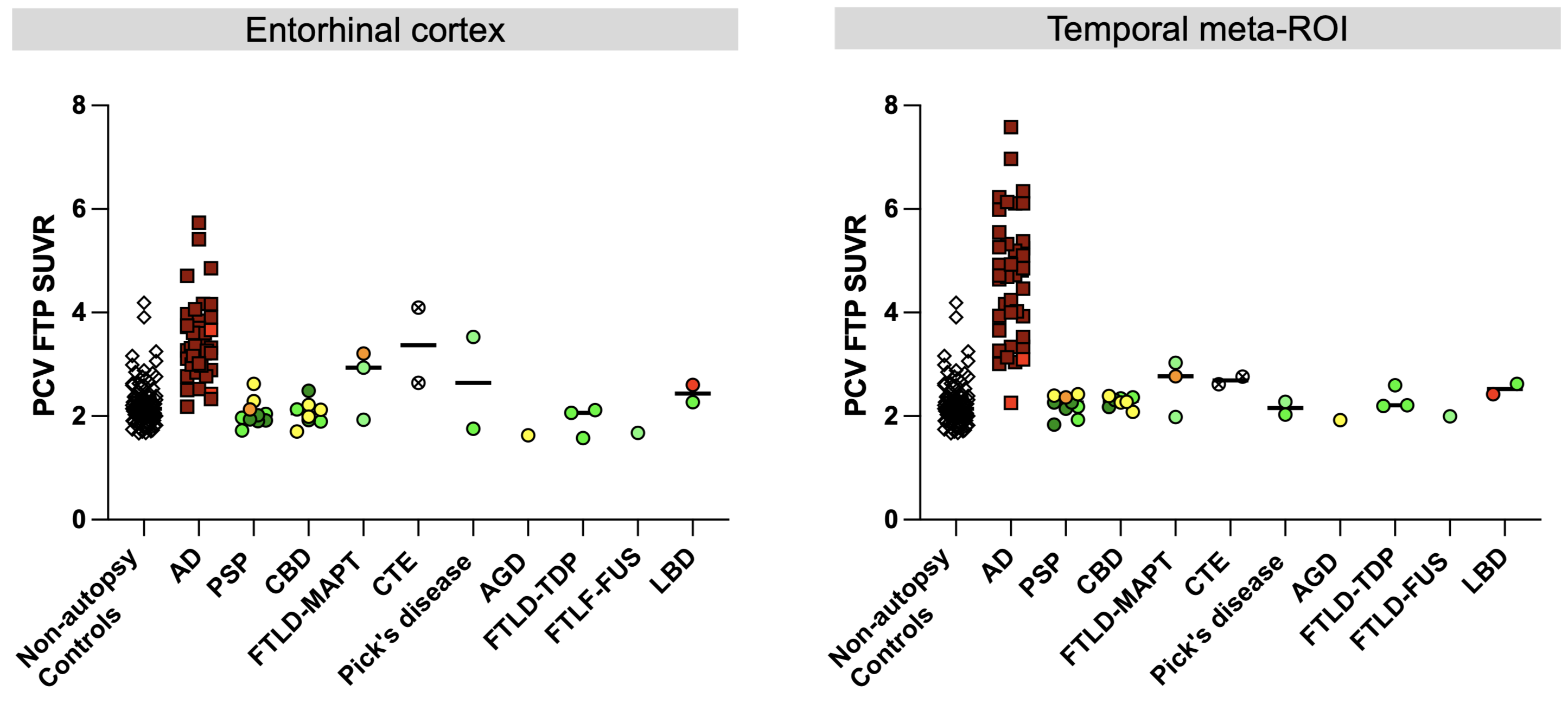

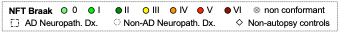

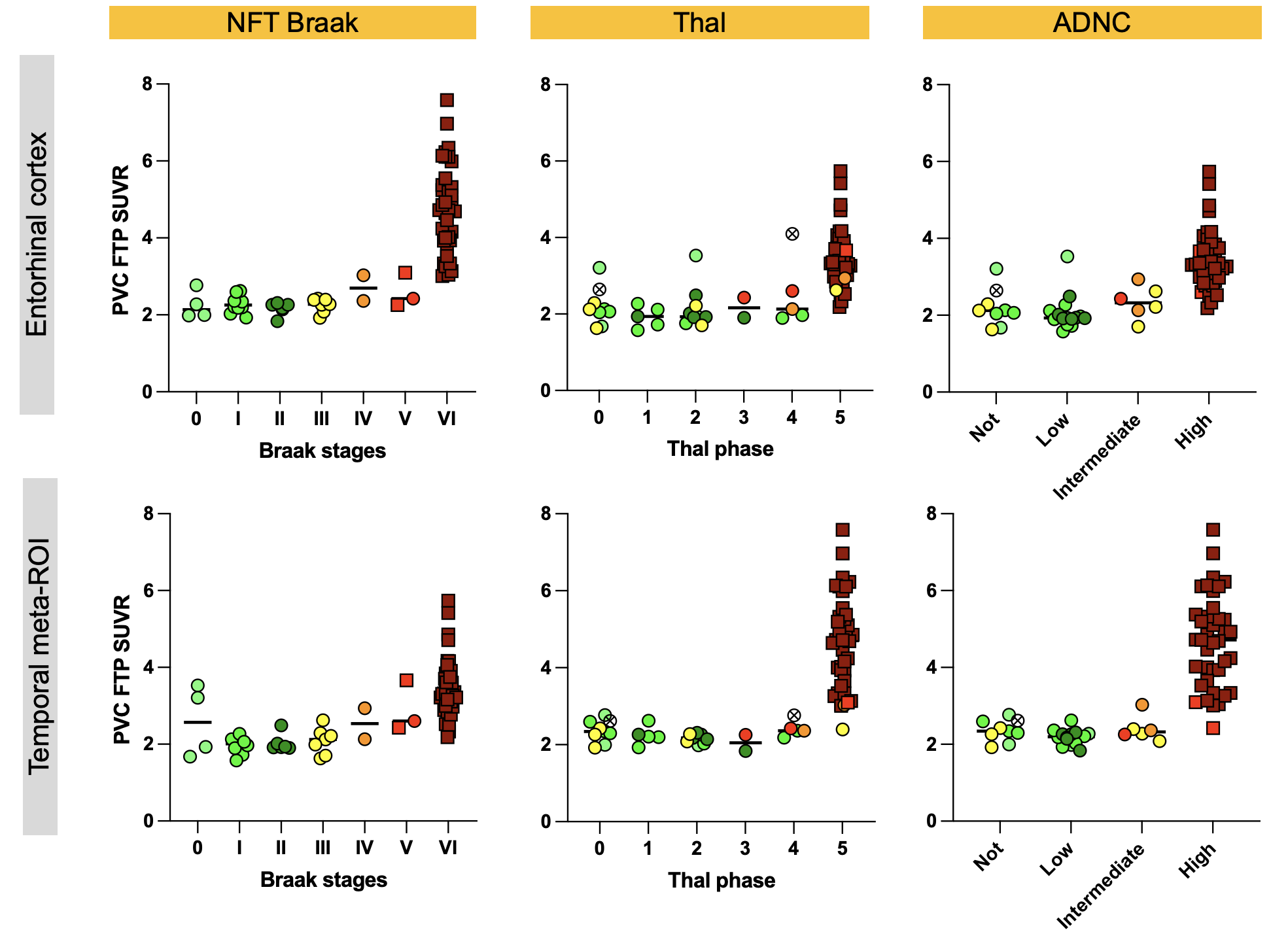
 **Supplementary Figure 4, Partial volume corrected Flortaucipir SUVRs across diagnoses and neuropathology scores**

**A,** Flortaucipir PVC SUVRs in the entorhinal cortex and the temporal meta-ROI by diagnosis

**B,** A Flortaucipir PVC SUVRs by NFT Braak stages, Thal phases, and ADNC levels.

PVC was performed using the PET PVC toolbox in n=71 participants (Thomas *et al.*, Physics in Medicine and Biology 2016). Two patients failed the PVC pipeline (n=1 CBD and n=1 PSP).

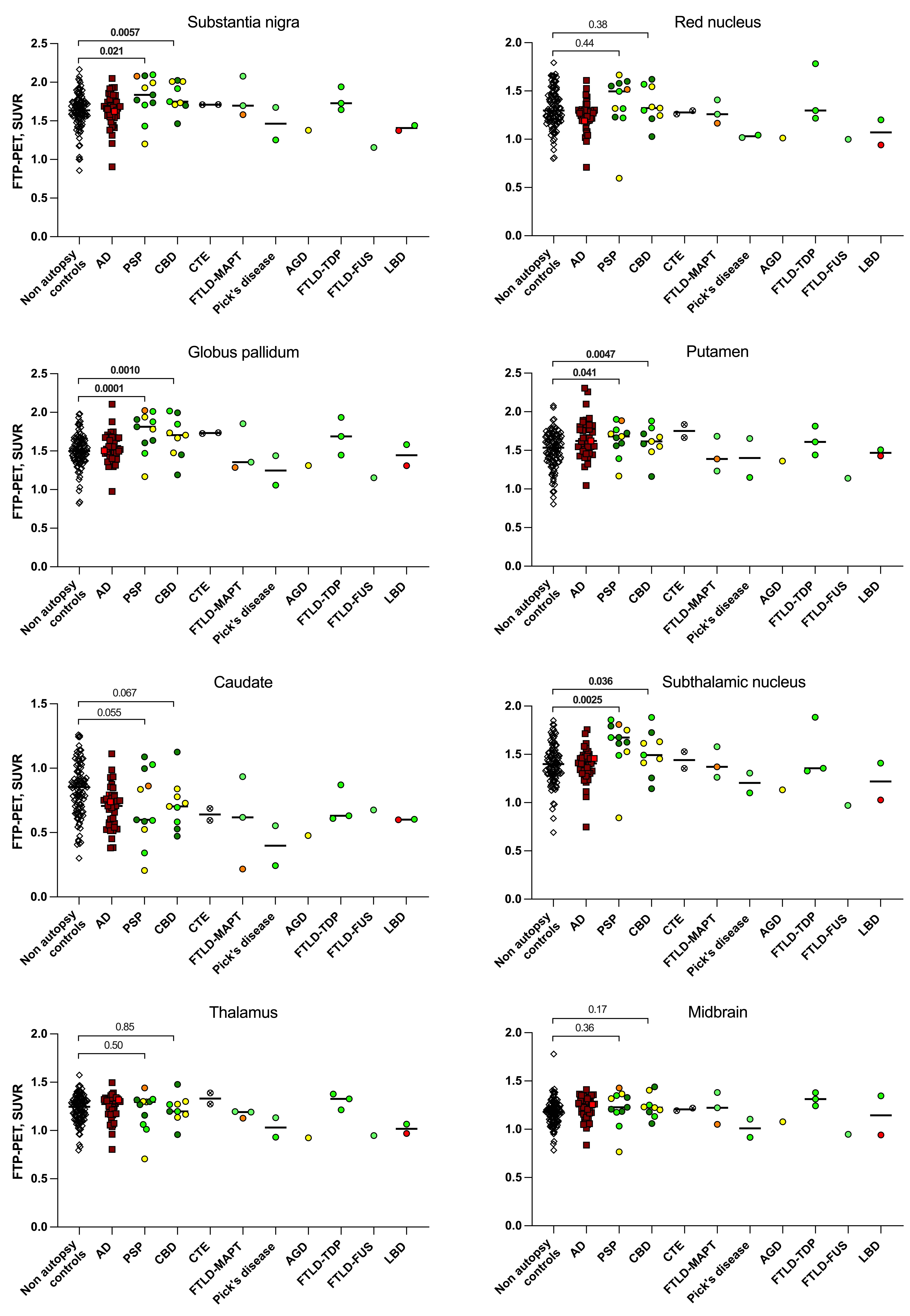

**Supplementary Figure 5, Flortaucipir SUVRs in basal ganglia across diagnosis groups**

We compared the PSP and CBD groups with controls as patterns of basal ganglia uptake have been reported in the literature in primary tauopathies. Other primary tauopathy diagnoses were not compared with controls as they included fewer than 5 cases. Comparisons were performed using linear regression adjusting for age.

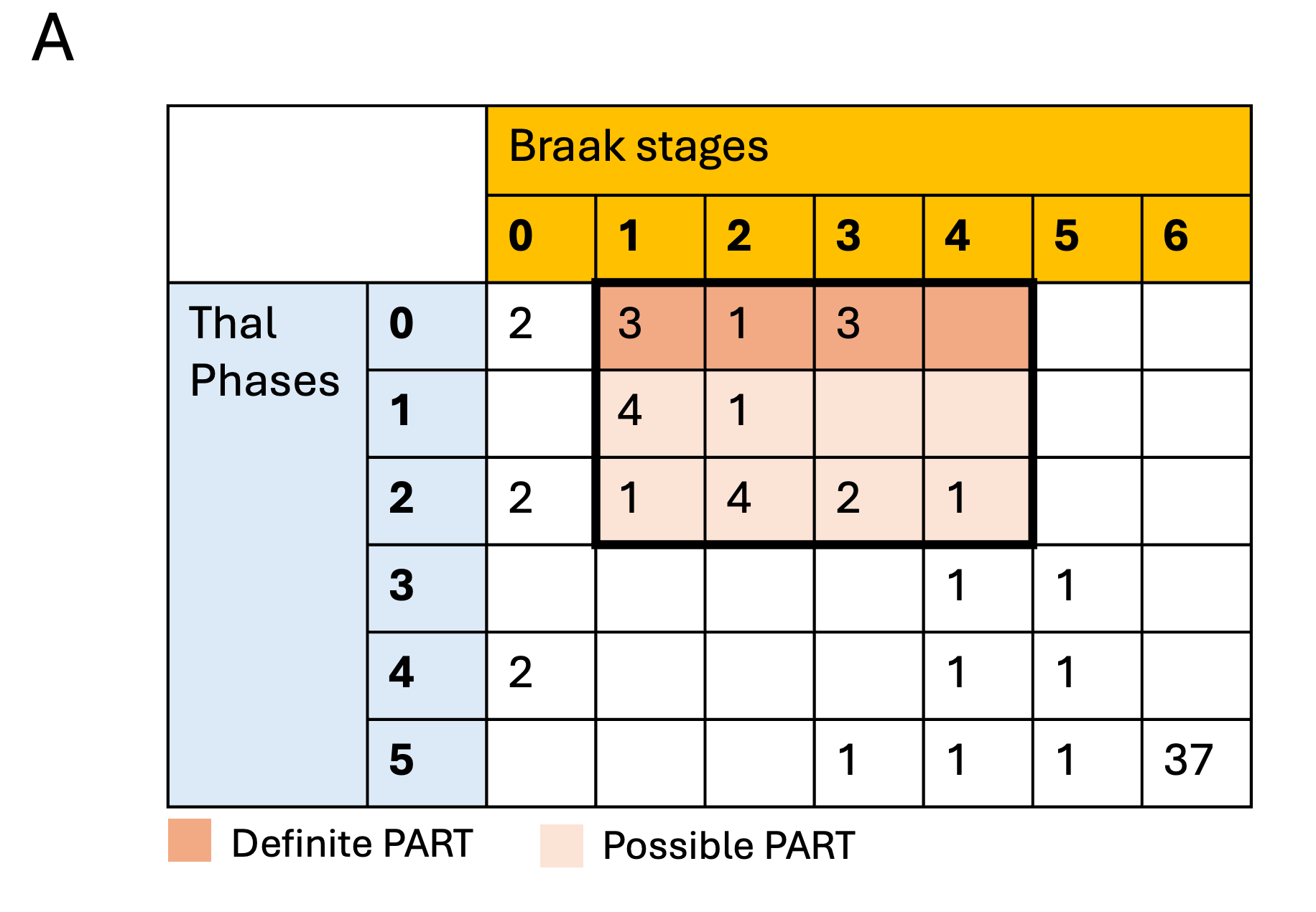

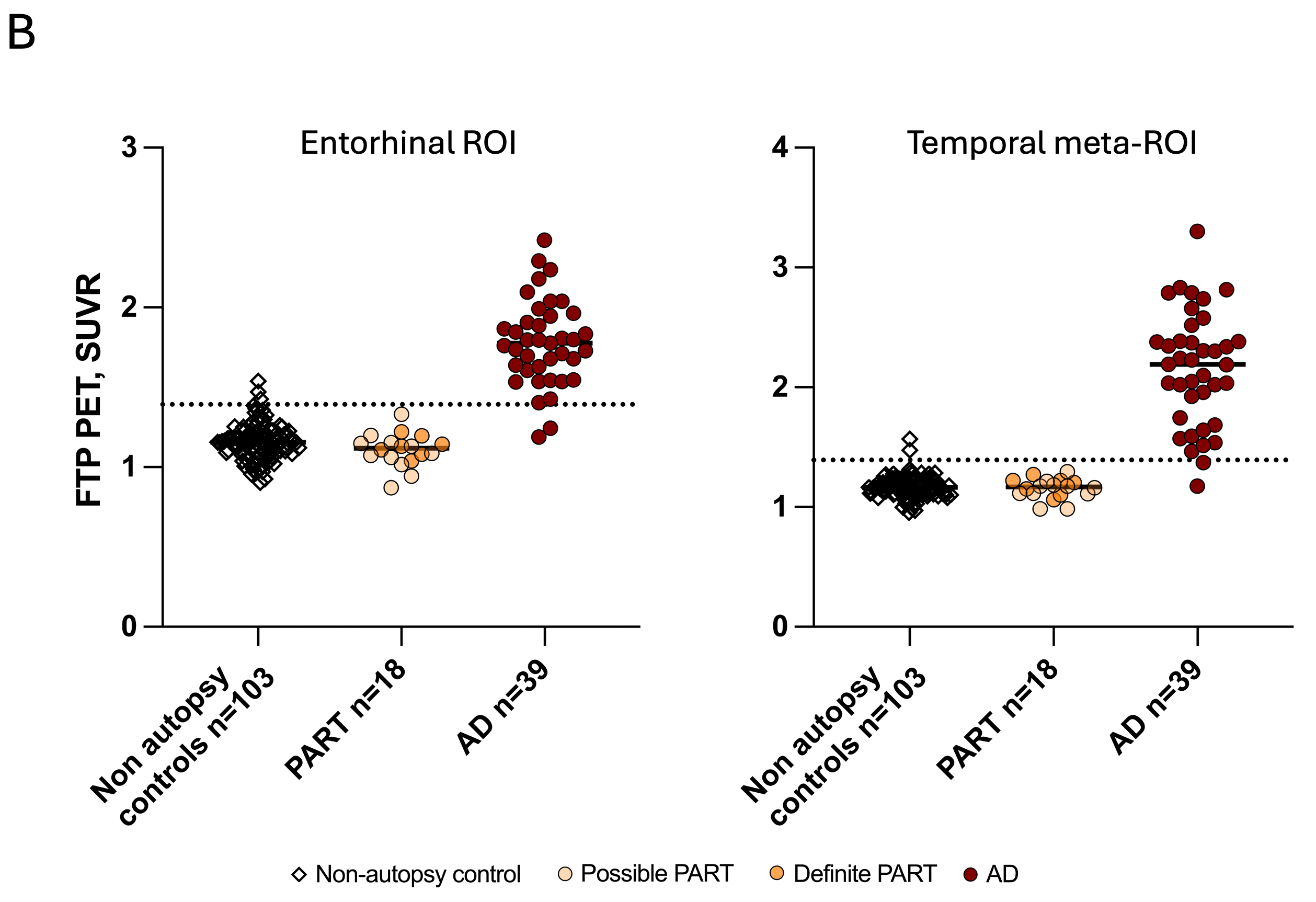

**Supplementary Figure 6, Flortaucipir SUVRs in PART and low-amyloid/Braak positive FTLD-tau cases**

**A,** Cohort cases by Braak and Thal stages and PART/PART-like cases. PART cases included n=3 FTLD-TDP cases and n=1 LBD. Low amyloid/ positive Braak cases (n=14) included FTLD-tau cases: AGD n=1, CBD n=6, PSP n=6, Pick’s n=1. Not included in the table: n=2 CTE cases Braak nonconformant; n=1 CBD with undetermined Thal phase.

**B,** FTP SUVR in the entorhinal cortex and temporal meta-ROI in PART cases, controls, and AD cases

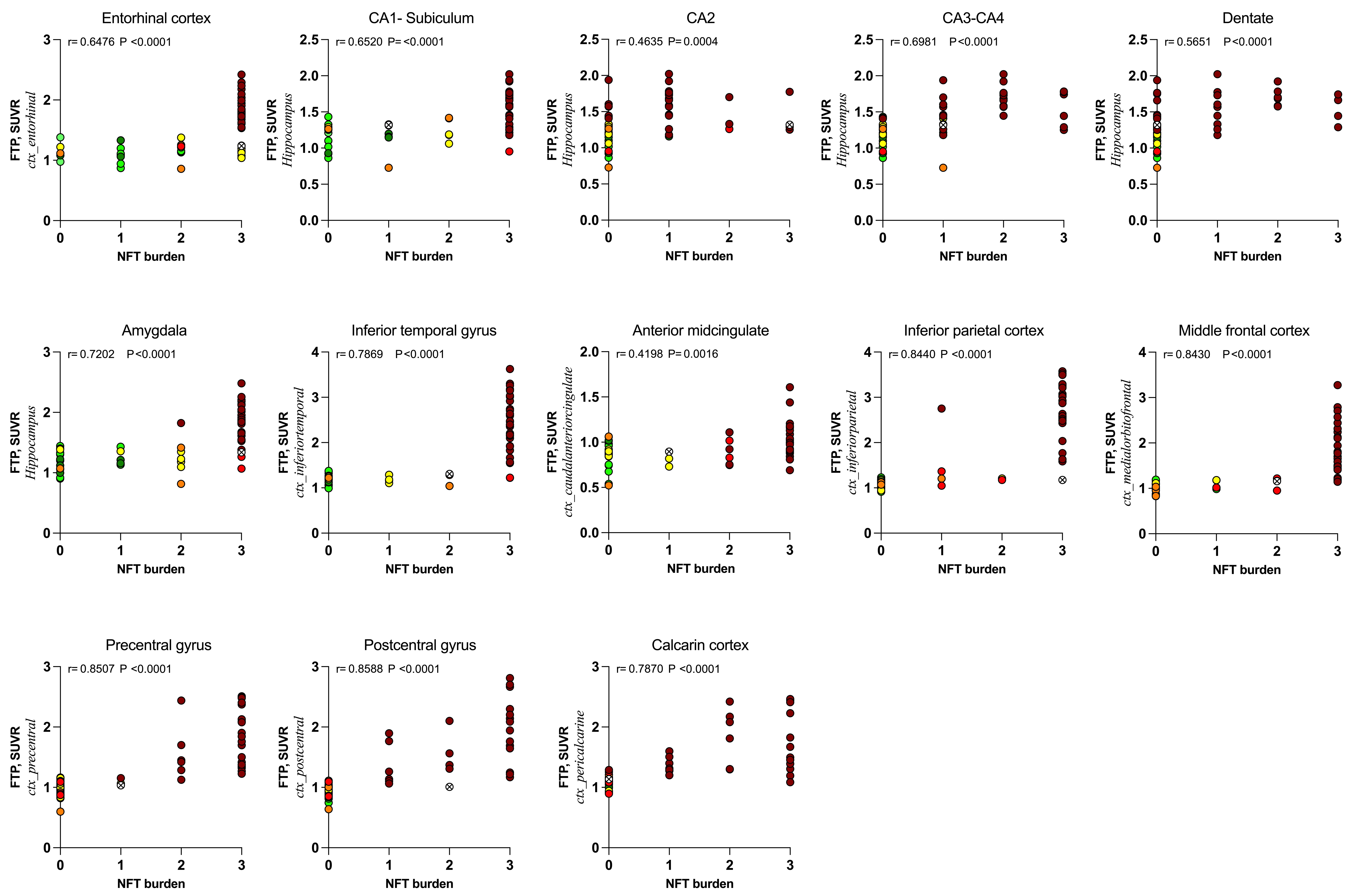

**Supplementary Figure 7, Association of AD NFT burden with Flortaucipir SUVRs across cortical regions**

Association of FTP-SUVR with AD NFT burden across the different brain regions. Correlations were analyzed with Spearman's rank correlations (r). Freesurfer labels of the PET ROI used for correlation are indicated in italics. n=56.

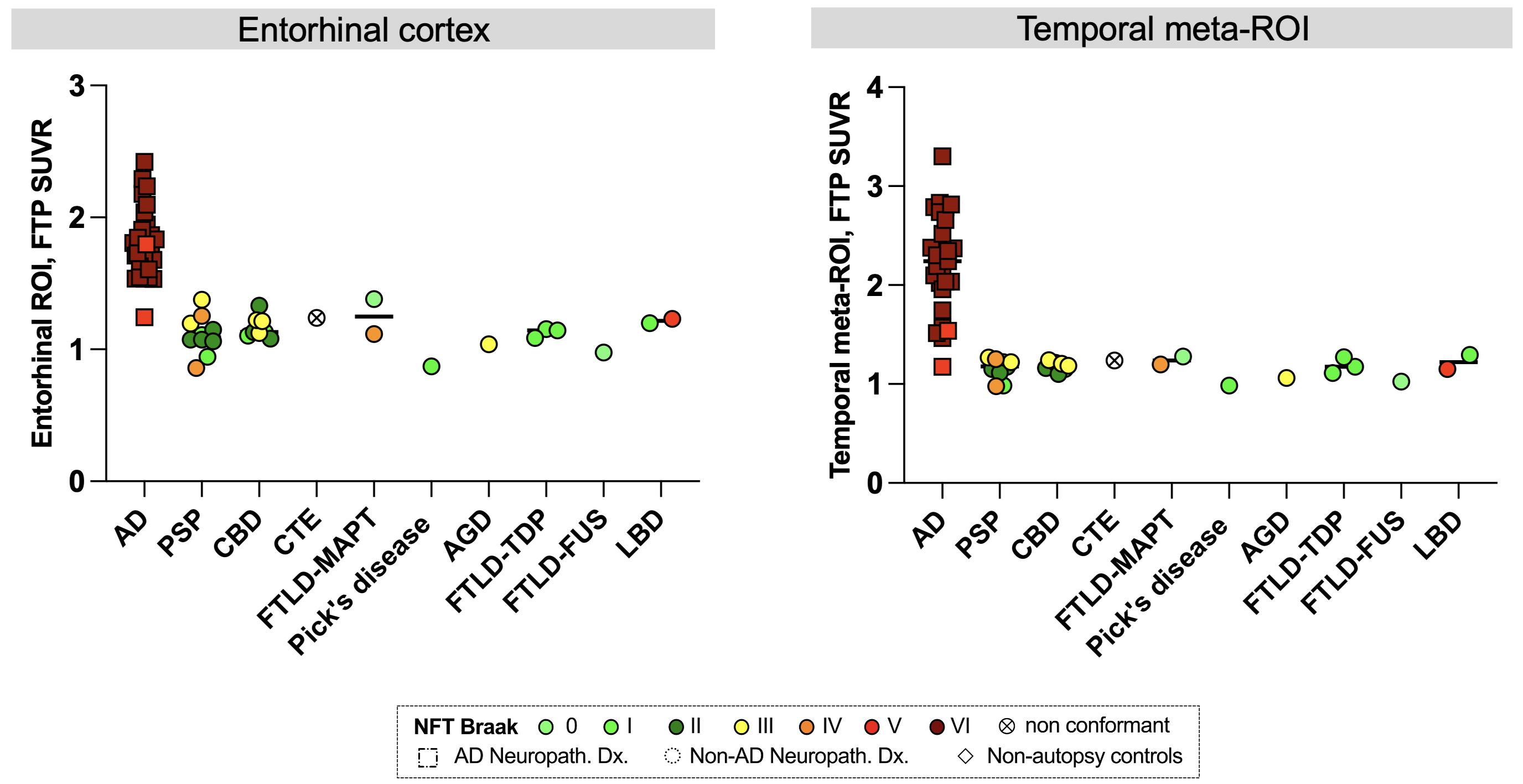

**B**

**A**

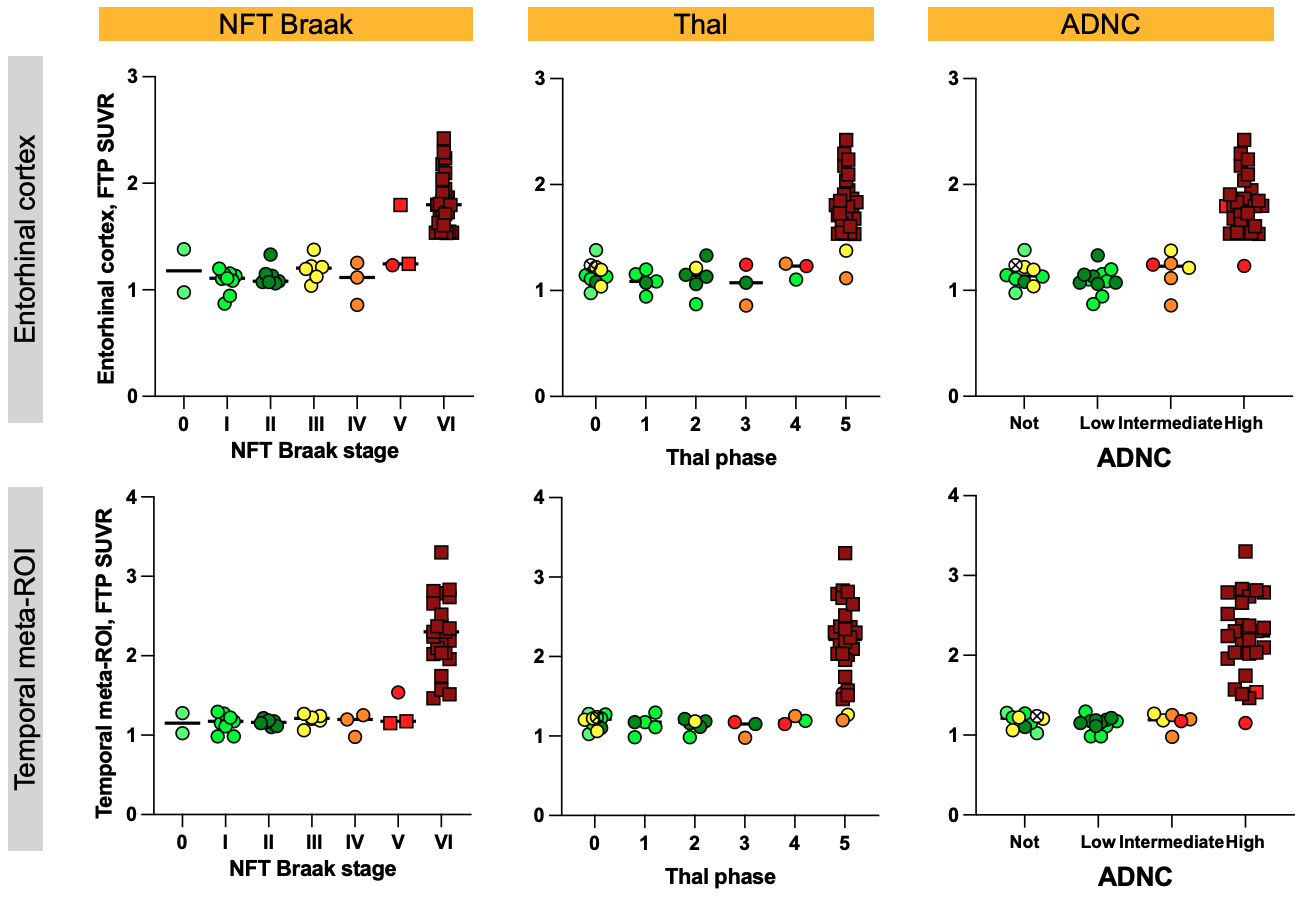

**Supplementary Figure 8, Flortaucipir-PET SUVRs across diagnoses and association with neuro pathological scores in the plasma subsample**

**A,** Flortaucipir SUVR in entorhinal and meta-temporal ROI by diagnosis. **B,** Flortaucipir SUVR in entorhinal and meta-temporal ROIs by Braak stages, Thal phases, and ADNC levels.

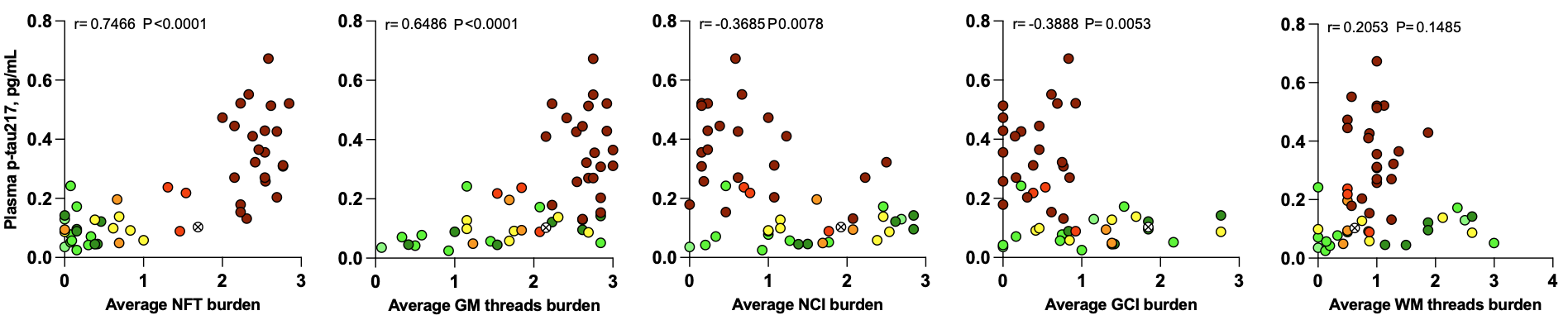

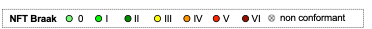

**Supplementary Figure 9, Association of plasma p-tau217 levels with AD NFT burden**

P-tau217 levels versus the average burden of NFT, across all brain cortical regions. Correlation was analyzed with Spearman's rank correlations (r). n=51.

|  | Overall | | AD | PSP | | CBD | | CTE | | FTLD-MAPT | | Pick's | | AGD | | FTLD-TDP | | FTLD-FUS | | LBD |
| --- | --- | --- | --- | --- | --- | --- | --- | --- | --- | --- | --- | --- | --- | --- | --- | --- | --- | --- | --- | --- |
|  | **(N=56)** | | **(N=27)** | **(N=10)** | | **(N=8)** | | **(N=1)** | | **(N=2)** | | **(N=1)** | | **(N=1)** | | **(N=3)** | | **(N=1)** | | **(N=2)** |
| Age at PET | 67.6 [34.4, 96.7] | | 67.1 [49.7, 96.7] | 72.6 [60.7, 81.3] | | 63.4 [47.3, 80.4] | | 66-70 | | 31-40, 66-70 | | 61-65 | | 66-70 | | 53.9 [48.3, 68.1] | | 31-40 | | 71-75, 81-85 |
| Sex, male | 33 (59%) | | 12 (44%) | 8 (80%) | | 5 (62%) | | 1 (100%) | | 1 (50%) | | 1 (100%) | | 1 (100%) | | 1 (33%) | | 1 (100%) | | 2 (100%) |
| ApoE4 carriership | 25/52 (48%) | | 17/26 (65%) | 3/7 (43%) | | 2 (25.0%) | | 0/1 (0%) | | 1/2 (50.0%) | | 1/2 (100%) | | 0/1 (0%) | | 1/3 (33%) | | 0/1 (0%) | | 0/2 (0%) |
| MMSE | 22 [0, 30] | | 19 [8, 27] | 26 [18, 29] | | 24 [9, 30] | | 27 | | 4 [4, 5] | | 25 | | 29 | | 24 [21, 26] | | 22 | | 14 [0, 28] |
| Plasma p-tau217 | 0.155  [0.0251, 0.673] | | 0.312  [0.0890, 0.673] | 0.0673  [0.0370, 0.197] | | 0.130  [0.0585, 0.172] | | 0.103 | | 0.112  [0.0940, 0.129] | | 0.0517 | | 0.0993 | | 0.0423  [0.025, 0.071] | | 0.035 | | 0.240  [0.238, 0.242] |
| Age at autopsy, years | 70.0  [35.4, 98.0] | | 70.7  [52.0, 98.0] | 75.4  [61.7, 84.9] | | 64.6  [49.9, 82.4] | | 71-75 | | 31-40, 66-70 | | 66-70 | | 66-70 | | 58.2  [50.0, 69.6] | | 31-40 | | 71-75, 86-90 |
| Plasma to PET delay, months | 1.7 [0.5, 4.1] | | 0.5 [0.4-4.1] | 1.9 [0.13, 9.5] | | 0.6 [0,2.9] | | 2.3 | | 0, 3.9 | | 1.2 | | 0.26 | | 1.7 [0, 1.8] | | 3.9 | | 0, 1.2 |
| Plasma-to-autopsy delay, years | 4.3 [3.3, 5.3] | | 4.3 [3.3, 5.3] | 3.2 [1.8, 4.1] | | 2.6 [1.8, 4.3] | | 4.1 | | 3.0, 2.5 | | 3.1 | | 0.34 | | 1.7 [1.6, 4.4] | | 1.3 | | 1.9, 3.0 |
| Thal phase |  | |  |  | |  | |  | |  | |  | |  | |  | |  | |  |
| 0 | 10 (18%) | | - | 2 (20%) | | 3 (37%) | | 1 (100%) | | 1 (50%) | | - | | 1 (100%) | | 1 (33%) | | 1 (100%) | | - |
| 1 | 5 (9%) | | - | 2 (20%) | | - | | - | | - | | - | | - | | 2 (67%) | | - | | 1 (50%) |
| 2 | 6 (11%) | | - | 2 (20%) | | 3 (37%) | | - | | - | | 1 (100%) | | - | | - | | - | | - |
| 3 | 3 (5%) | | 1 (4%) | 2 (20%) | | - | | - | | - | | - | | - | | - | | - | | - |
| 4 | 3 (5%) | | - | 1 (10%) | | 1 (12%) | | - | | - | | - | | - | | - | | - | | 1 (50%) |
| 5 | 28 (50%) | | 26 (97%) | 1 (10%) | | - | | - | | 1 (50%) | | - | | - | | - | | - | | - |
| Unstagable | 1 (2%) | | - | - | | 1 (12%) | | - | | - | | - | | - | | - | | - | | - |
| NFT Braak stage | |  | | |  | |  | |  | |  | |  | |  | |  | |  | |
| 0 | 2 (4%) | | - | - | | - | | - | | 1 (50%) | | - | | - | | - | | 1 (100%) | | - |
| I | 9 (16%) | | - | 2 (20%) | | 2 (25%) | | - | | - | | 1 (100%) | | - | | 3 (100%) | | - | | 1 (50%) |
| II | 7 (12%) | | - | 4 (40%) | | 3 (37%) | | - | | - | | - | | - | | - | | - | | - |
| III | 6 (11%) | | - | 2 (20%) | | 3 (37%) | | - | | - | | - | | 1 (100%) | | - | | - | | - |
| IV | 3 (5%) | | - | 2 (20%) | | - | | - | | 1 (50%) | | - | | - | | - | | - | | - |
| V | 3 (5%) | | 2 (7%) | - | | - | | - | | - | | - | | - | | - | | - | | 1 (50%) |
| VI | 25 (45%) | | 25 (93%) | - | | - | | - | | - | | - | | - | | - | | - | | - |
| Non conformant | 1 (2%) | | - | - | | - | | 1 (100%) | | - | | - | | - | | - | | - | | - |
| AD CERAD |  | |  |  | |  | |  | |  | |  | |  | |  | |  | |  |
| None | 16 (29%) | | - | 5 (50%) | | 3 (37.5%) | | 1 (100%) | | 1 (50%) | | - | | 1 (100%) | | 3 (100%) | | 1 (100%) | | 1 (50%) |
| Sparse | 3 (5%) | | - | 1 (10%) | | 1 (12.5%) | | - | | - | | 1 (100%) | | - | | - | | - | | - |
| Moderate | 8 (14%) | | - | 4 (40%) | | 3 (37.5%) | | - | | 1 (50%) | | - | | - | | - | | - | | - |
| Frequent | 29 (52%) | | 27 (100%) | - | | 1 (12.5%) | | - | | - | | - | | - | | - | | - | | 1 (50%) |
| ADNC |  | |  |  | |  | |  | |  | |  | |  | |  | |  | |  |
| Not | 10 (18%) | | - | 2 (20%) | | 3 (37%) | | 1 (100%) | | 1 (50%) | | - | | 1 (100%) | | 1 (33%) | | 1 (100%) | | - |
| Low | 12 (22%) | | - | 1 (50%) | | 3 (37%) | | - | | - | | 1 (100%) | | - | | 2 (67%) | | - | | 1 (50%) |
| Intermediate | 6 (11%) | | 1 (4%) | 3 (30%) | | 1 (12%) | | - | | 1 (50%) | | - | | - | | - | | - | | - |
| High | 27 (48%) | | 26 (96%) | - | | - | | - | | - | | - | | - | | - | | - | | 1 (50%) |
| Unstagable | 1 (2%) | | - | - | | 1 (12%) | | - | | - | | - | | - | | - | | - | | - |
| Not | 10 (18%) | | - | 2 (20%) | | 3 (37%) | | 1 (100%) | | 1 (50%) | | - | | 1 (100%) | | 1 (33%) | | 1 (100%) | | - |

**Supplementary Table 1, Demographics and plasma biomarkers levels in the plasma subsample (n=56).**

Categorical variables are presented as median [Q1, Q3] for groups of n≥3. Individual values are presented for groups of n=1-2. APOE4 carriership is presented as the number of carriers/number of cases tested (% of positivity). To limit identifying information, ages are presented as age range for diagnosis group of n=1 to 2.

Abbreviations: ADNC, Alzheimer's disease neuropathologic change; AGD, argyrophilic grain disease; CBD, corticobasal degeneration; CERAD, Consortium to Establish a Registry for Alzheimer's Disease; CTE, chronic traumatic encephalopathy; FTLD, frontotemporal lobar degeneration; Fus, RNA-binding protein fused in sarcoma; LBD, Lewy body dementia; MMSE, mini-mental state examination; NFT, neurofibrillary tangles; PSP, progressive supranuclear palsy.
